# SPIRIT-CONSORT-ELM: Element-Level Annotated Dataset and Large Language Model Approach for Assessing Randomized Controlled Trial Reporting

**DOI:** 10.64898/2026.06.06.26354746

**Authors:** Lan Jiang, Xiangji Ying, Andrew W. Brown, Mengfei Lan, Wenxuan Song, Joe D. Menke, Colby J. Vorland, Evan Mayo-Wilson, Halil Kilicoglu

**Affiliations:** School of Information Sciences, University of Illinois Urbana-Champaign, IL, USA; Department of Epidemiology, University of North Carolina Gillings School of Global Public Health, Chapel Hill, NC, USA; Department of Biostatistics, University of Arkansas for Medical Sciences, Little Rock, AR, USA; Arkansas Children’s Research Institute, Little Rock, AR, USA; Department of Epidemiology and Biostatistics, Indiana University School of Public Health-Bloomington, Bloomington, IN, USA

## Abstract

Randomized controlled trials (RCTs) are central to assessing the benefits and harms of interventions, but incomplete reporting undermines their verifiability and usefulness. Although SPIRIT and CONSORT reporting guidelines promote complete reporting of RCT protocols and results publications, many RCTs remain incompletely reported. Automated manuscript checking could help improve reporting completeness before publication. We previously developed SPIRIT-CONSORT-TM, a corpus of 200 articles (100 protocol-results publication pairs) annotated with 83 checklist items from SPIRIT 2013 and CONSORT 2010, and trained models for item-level assessment. However, checklist items may comprise multiple constituent elements, which prior work did not capture or evaluate. Here, we extend the corpus with element-level annotations (SPIRIT-CONSORT-ELM) and formulate assessment as a machine reading comprehension task operationalized through 119 questions targeting specific reporting elements. Two annotators independently assessed 50 articles (25 pairs), with discrepancies resolved through discussion; one annotator assessed the remaining 150 articles. We then developed an automated pipeline combining PubMedBERT-based evidence retrieval with GPT-5-based question answering. Inter-annotator agreement was high (Gwet’s AC1: 0.782), and the pipeline achieved high performance (F1: 0.822, Gwet’s AC1: 0.796). Component analyses demonstrated the importance of evidence retrieval quality and modest benefits from illustrative in-context examples. SPIRIT-CONSORT-ELM provides a benchmark for fine-grained assessment of RCT reporting completeness, while the automated pipeline establishes a robust baseline and shows potential for supporting authors, reviewers, and editors.

## Introduction

Randomized controlled trials (RCTs) provide important evidence about the benefits and harms of therapeutic interventions.^1^ The verifiability and usability of RCTs depend on complete and accurate reporting of study design, conduct, analyses, and results. However, RCT reporting is frequently incomplete.^2–5^ Incomplete reporting can hinder proper interpretation of results, and contribute to research waste.^6^ The selective non-reporting of studies and results can also lead to bias in the literature overall, as in systematic reviews and meta-analyses that include “positive” results but miss “negative” or “null” results that were never reported. To fully realize their potential benefits for clinical practice and health policy, RCTs must be reported completely. Two reporting guidelines aim to help authors report RCT protocols and results. SPIRIT (Standard Protocol Items: Recommendations for Interventional Trials) provides guidance for trial protocols.^7,8^ CONSORT (Consolidated Standards of Reporting Trials) focuses on reporting trial results.^9,10^ Each guideline comprises a checklist and flow diagram, specifying the minimum information recommended for inclusion. Both guidelines were updated in 2025.^8,10^

Although CONSORT and SPIRIT are widely endorsed by journals and other organizations, many RCTs are not reported completely. Journal endorsement is not always accompanied by procedures to verify that reports include the recommended information.^11^ Evaluating RCT submissions before peer review improves reporting completeness; however, manual assessment is time-consuming, requires substantial expertise, and is infeasible for all but a small number of well-resourced journals.^3,12,13^ This limitation has motivated meta-research aimed at evaluating reporting completeness; however, existing studies use diverse approaches to derive item-level judgments and quantify adherence to reporting guidelines.^14^ In addition, many authors may be unaware of reporting guidelines or lack sufficient guidance on how to comply with them. These challenges are exacerbated by the growing volume of manuscript submissions, highlighting the need for scalable and consistent approaches to screening reporting completeness.^15–17^ Natural language processing (NLP) and artificial intelligence (AI) offer promising solutions to checking whether manuscripts are complete and to recommend improvements before articles are published. In recent years, they have been applied to improve reporting completeness using established reporting guidelines. Early work introduced CONSORT-TM, an annotated corpus of 50 RCT results publications labeled with CONSORT 2010 checklist items, and investigated BERT-based fine-tuning, as well as data augmentation, weak supervision, and BioGPT fine-tuning to identify sentences reporting these items.^18–20^ Using these models, they also conducted large-scale analyses of historical reporting trends, confirming that while some items (e.g., outcome definitions) were commonly reported, others (e.g., allocation concealment) remained infrequently reported.^21^ Subsequent work expanded this effort through the development of SPIRIT-CONSORT-TM, a corpus comprising 100 paired RCT protocols and results publications annotated with a unified set of 83 checklist items drawn from SPIRIT and CONSORT.^22^ An NLP model trained on this dataset achieved F_1_ scores of 0.74 at sentence level (SPIRIT: 0.75; CONSORT: 0.75) and 0.87 at the article level (SPIRIT: 0.89; CONSORT: 0.92). More recently, generative large language models (LLMs) have been applied to these corpora for checking article-level completeness, yielding comparable performance.^17,23^ Beyond RCT publications, LLMs have also been applied to evaluate compliance of systematic reviews with PRISMA 2009 guidelines, achieving performance around 0.70, with hybrid human-AI approaches reaching accuracies of 0.89-0.96.^24^

Previous research largely frames reporting completeness as a binary determination of whether each checklist item is present in a publication or not. However, most SPIRIT and CONSORT checklist items comprise multiple distinct elements.^1,25^ For example, SPIRIT 2013 item 30 (Ancillary and post-trial care) recommends that authors describe ancillary and post-trial care plans, compensation for trial-related harm, or provide a rationale if such plans are absent. In practice, authors may only partially report this information. Existing corpora and models typically check reporting completeness at the item level (i.e., determining whether any content related to a checklist item is present) without checking its constituent elements individually. This approach can misrepresent reporting completeness by marking items as complete despite missing key elements, or as incomplete without distinguishing whether a single element or multiple elements are absent. Moreover, it is often unclear what criteria the models use to make judgments, particularly for vague and compound checklist items. Checking element-level RCT reporting completeness is therefore needed to enable valid and reliable evaluation of reporting completeness. Such fine-grained assessment may also improve comparability across studies, as previous estimates of reporting completeness often differ because studies evaluate different subsets of checklist items and apply different definitions of what constitutes “complete” reporting. ^3,14^

In this study, we assessed the completeness of RCT publications at the element level. To enable this analysis, we developed a comprehensive element-level reporting completeness framework, curated a dataset, and investigated automated approaches to assess the completeness of RCT publications at the element level.

The primary contributions of this work are threefold:

1. *Element-level reporting completeness framework*: We formalized 119 questions with corresponding answer choices derived from 83 items in SPIRIT 2013 and CONSORT 2010. Each question targets specific elements within a checklist item, enabling fine-grained assessment of reporting beyond binary item-level presence.
2. *Element-level annotated dataset*: We curated SPIRIT-CONSORT-ELM (Element), an expert-annotated dataset in which 119 questions were answered for 200 articles (100 protocol-results pairs) from SPIRIT-CONSORT-TM.^22^ We also made this dataset publicly available.
3. *LLMs for element-level RCT completeness assessment*: We evaluated the feasibility of automating element-level assessment by prompting two LLMs (proprietary GPT-5^26^ and open-weight Qwen-2.5^27^) to answer each question using item-related text annotated by experts or extracted by prior models.^22^ Model performance was validated against expert annotations using F_1_ score, sensitivity, specificity, and Gwet’s AC1.

Together, SPIRIT-CONSORT-ELM and the accompanying evaluations establish a benchmark for fine-grained, element-level assessment of RCT reporting completeness, providing a foundational resource for developing and validating automated systems to improve reporting.

## Results

In this section, we present the results of dataset curation and automated element-level reporting assessment. We first summarize the characteristics of the curated dataset, including inter-annotator agreement and question distribution by type and reporting guideline applicability. We then evaluate an LLM-based framework for assessing reporting completeness by formulating each each of the 119 questions as a machine reading comprehension task, in which the model answers the question based on relevant article text. We compare several assessment pipelines, examining the effects of LLM backbones, evidence snippet identification methods, and strategies for selecting examples for prompting.

### Descriptive statistics for SPIRIT-CONSORT-ELM

Table 1 provides high-level descriptive statistics for the SPIRIT-CONSORT-ELM dataset. Of 119 questions, 85 (71%) are “check-one”and 34 (29%) are “check-all-that-apply” questions. Check-all-that-apply questions have an average of 1.63 options selected per question. Two questions are multi-element; one requires a single-choice response for each element, whereas the other allows multiple-choice responses. 26 questions are conditionally dependent on the responses to preceding questions.

**Table 1.**
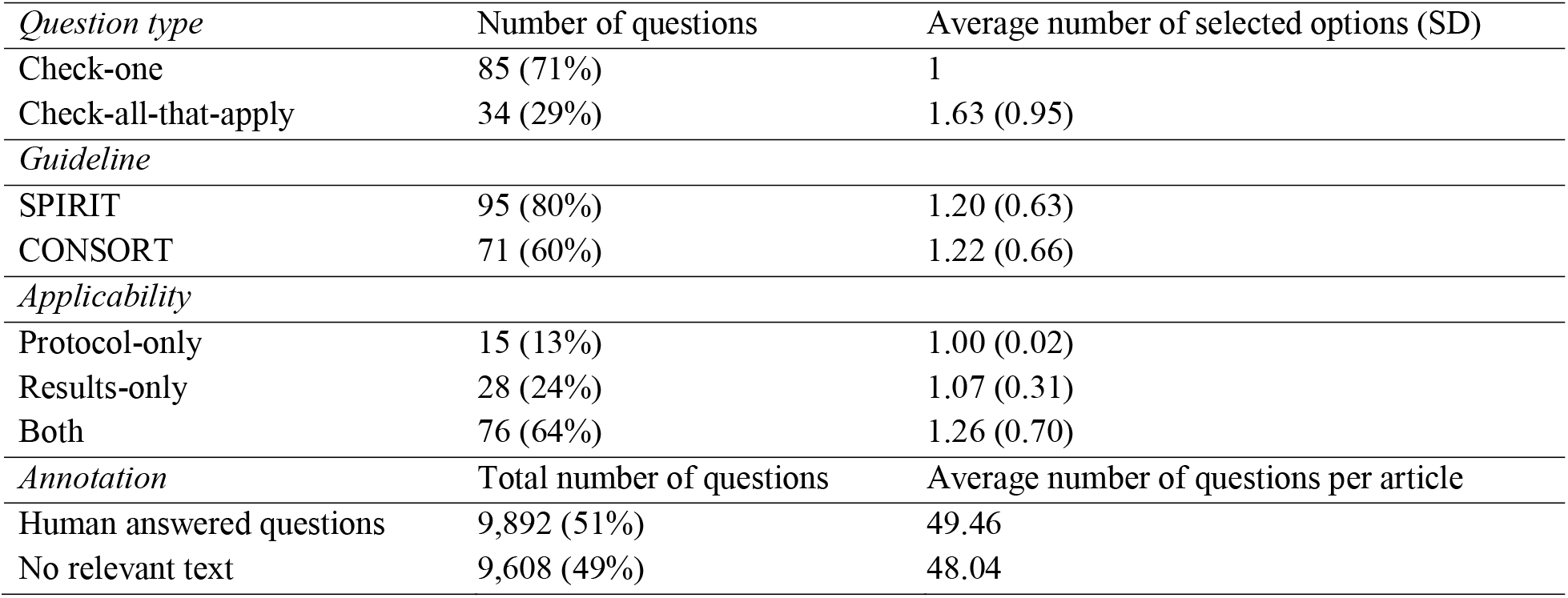
Descriptive statistics of question types and annotations in the SPIRIT-CONSORT-ELM dataset.

Of the 119 questions, 89 are associated with SPIRIT and 63 with CONSORT. 91 questions apply to protocols and 104 apply to results publications. The discrepancy between the number of questions associated with the report type and the corresponding guidelines reflects the fact that some questions derived from SPIRIT are also relevant to results publications, and some derived from CONSORT are applicable to protocols. Specifically, 76 questions (64%) apply to both protocol and results publications, whereas 15 questions (13%) are specific to protocols and 28 (24%) to results publications. Questions applicable to both report types have a higher number of selected response options on average (1.26 vs. 1.00 for protocols and 1.07 for results, respectively).

Across the dataset, annotators explicitly responded to 9,892 questions out of 19,500 potential questions (51%) (Table 1). For the remaining cases, either no relevant text was available for annotation or the question was not applicable. For example, the question about trial stopping (“Does the manuscript report that the trial was stopped early?”) applied to 13% of the results publications that reported early termination; the remaining 87% received automatic “No text to assess” responses. In the most extreme case, only seven articles (3.5%) reported code sharing and access, resulting in 96.5% of articles being automatically coded as “Not reported”. Overall, only one item (Participant inclusion criteria) was reported in all 200 articles.

For most questions, the responses tend to be highly skewed toward a single response, often “No” or “Not reported”, indicating that details to answer the question were missing, and in some cases, affirmative responses, indicating that related checklist items were well-reported. For example, for the question that asked whether an informed consent form was included as appendix, “No” response was selected for 198 articles (99%). In contrast, for the question assessing whether the manuscript specified where ethics approval has been obtained, or plans to seek such approval, “Yes” was selected for 194 articles (97%).

### Inter-annotator agreement

Inter-annotator agreement was assessed using 50 double-annotated articles (25 pairs), yielding a mean Gwet’s AC1 of 0.782 (SD=0.219) and a median of 0.821 (IQR=0.251). The two annotators achieved perfect agreement (Gwet’s AC1 = 1.00) on some questions, such as the question on whether the manuscript mentions where the protocol can be accessed (Box A in Methods below). Agreement was lower for questions requiring more interpretation. The lowest agreement (Gwet’s AC1 = 0.33) was observed on questions regarding whether adjusted analyses were pre-specified or conducted post hoc. When compared to the reconciled data, one annotator achieved a mean Gwet’s AC1 of 0.893 and the other achieved 0.891.

### Element-level reporting assessment using LLMs

The primary results are for the LLM pipeline that uses the existing SPIRIT-CONSORT-TM model for evidence retrieval, the hand-crafted prompt template for machine reading comprehension, and no in-context examples. The comparison of performance for a proprietary LLM (GPT-5) and an open-weight LLM (Qwen-2.5) is shown in Figure 1. In all evaluation metrics, both GPT-5 and Qwen-2.5 perform better than the majority class baseline, which simply assigns the answer most observed in the training set as the response.

**Figure 1.**
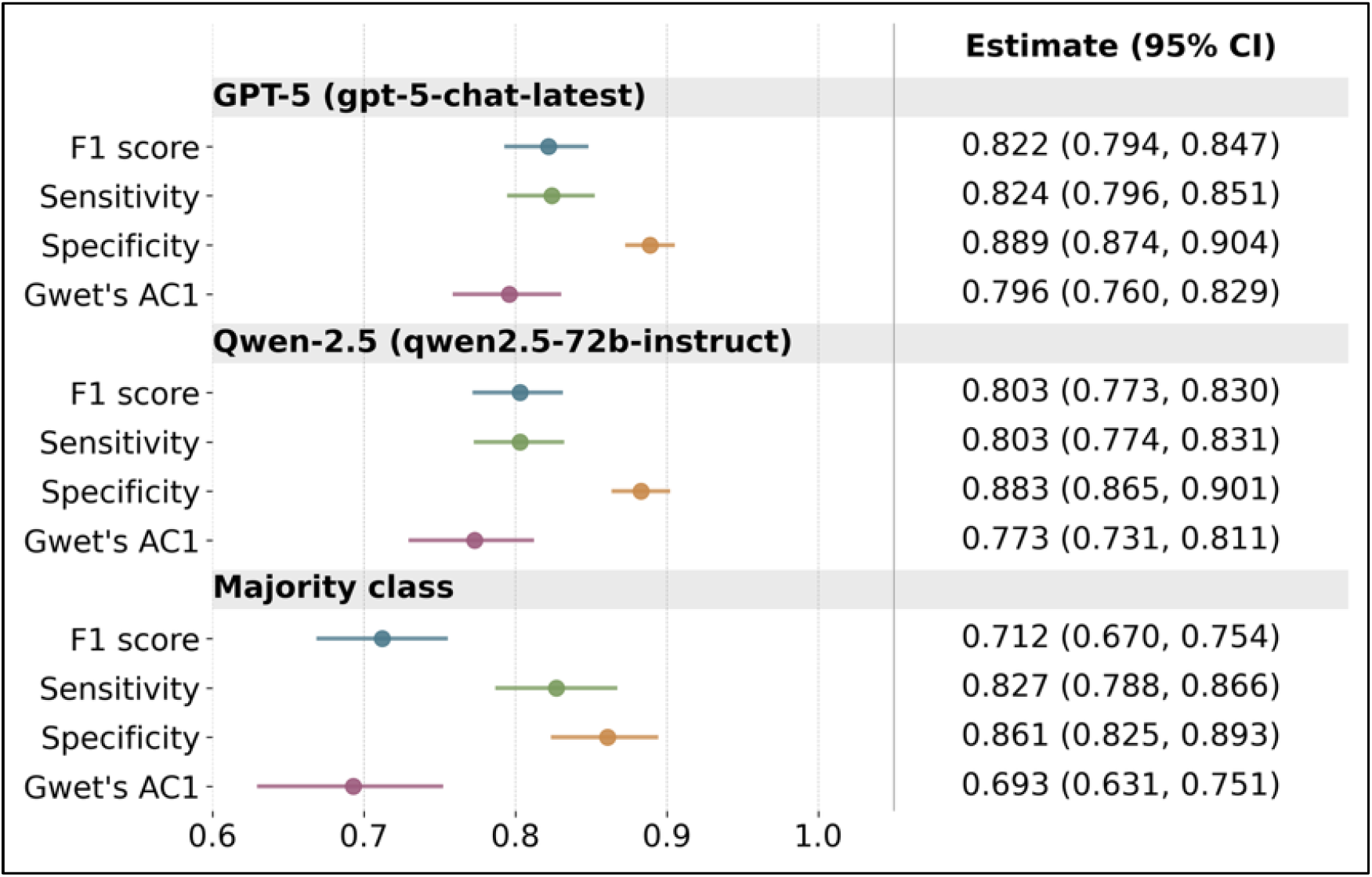
**Performance comparison of the LLM approaches and the majority class baseline**. F_1_, sensitivity, specificity, and Gwet’s AC1 scores are reported for 119 questions along with 95% CIs. LLM approaches use SPIRIT-CONSORT-TM-identified snippets and hand-crafted prompt template. CI: confidence interval.

GPT-5 achieves higher scores than Qwen-2.5 (+2.1 percentage points (pp) in F_1_, +2.1 pp in sensitivity, +0.6 pp in specificity, and +2.6 pp in Gwet’s AC1). Overall, GPT-5 and Qwen- 2.5 achieve comparable performance, with overlapping 95% CIs. As GPT-5 was the better-performing model, subsequent analyses focus on its results. Detailed question-level performance, including F_1_, Gwet’s AC1, sensitivity, specificity, PPV, and NPV, are reported in Supplementary File Section B. Analyses of model cost and computational efficiency are provided in Supplementary File Section D.

To assess the impact of evidence retrieval methods, we compared the use of SPIRIT-CONSORT-TM-based evidence retrieval with expert-annotated text. The results are summarized in Table 2. Expert-annotated snippets serve as an upper bound on performance. Using these snippets, GPT-5 achieves 0.893 F_1_, 0.894 sensitivity, 0.935 specificity, 0.877 Gwet’s AC1, higher than the results obtained with the default retrieval method (SPIRIT- CONSORT-TM model) reported above (+7.1 pp in F_1_, +7.0 pp in sensitivity, +4.6 pp in specificity and +8.1 pp in Gwet’s AC1). This suggests that further improving the quality of retrieved evidence can substantially enhance the performance of LLM-based element-level assessment.

**Table 2.**
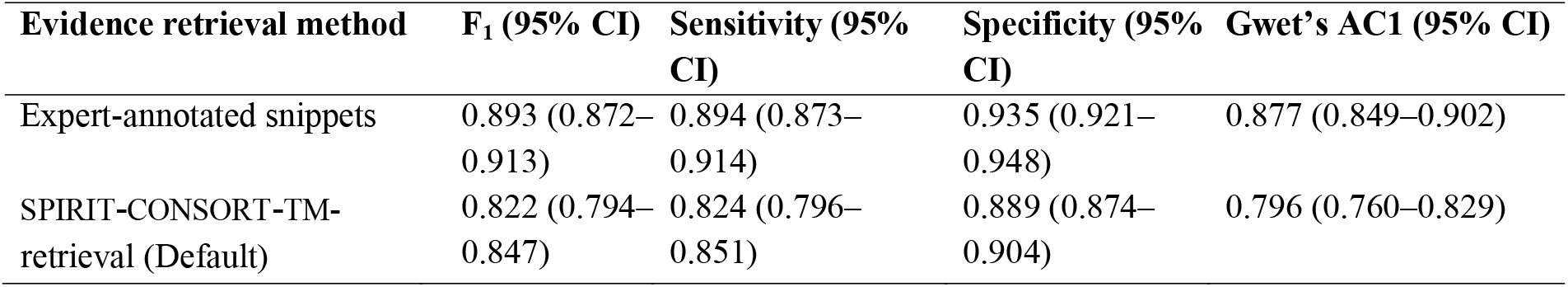
The comparison of impact of evidence retrieval methods on model performance. CI: confidence interval.

For the best-performing model, we also evaluated the impact of adding in-context examples. The results with the default evidence retrieval method (SPIRIT-CONSORT-TM) are shown in Table 3. Introducing a single example (one-shot) improves performance across all metrics, although the magnitude of improvement depends on the example selection strategy. Selecting examples based on semantic similarity using the Qwen-8B embedding model yields the best F_1_ and sensitivity (+0.8 pp, +0.6 pp, respectively, compared to the zero-shot setting), while random selection produces the highest specificity (+0.4 pp) and Gwet’s AC1 (+0.9 pp). Overall, however, differences across example selection strategies are minor.

**Table 3.**
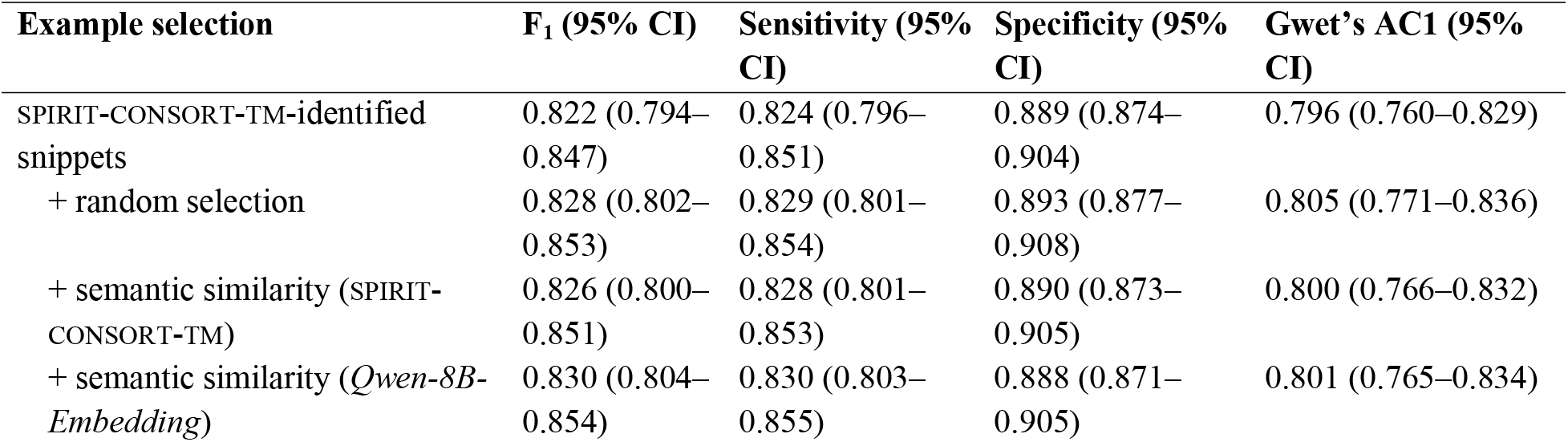
LLM performance on 119 questions using SPIRIT-CONSORT-TM–identified evidence under different example selection strategies. CI: confidence interval.

## Discussion

This study presents, to our knowledge, the first dataset and automated framework for assessing element-level reporting completeness in RCT publications. While the dataset is relatively modest in size in terms of number of publications included, it includes more than 19,000 judgments, providing sufficient scale for the development and evaluation of fine-grained assessment models. The dataset leverages and complements our prior work on item-level assessment.^22^

During the course of this study, updated SPIRIT 2025 and CONSORT 2025 guidelines were released. Comparison of our framework with these updated checklists showed that that 19 of 37 checklist items were fully represented and 18 were partially represented across 119 questions. For CONSORT 2025, 12 of 28 items were fully addressed and 16 were partially addressed. The updated guidelines further reinforce the need for fine-grained assessment, as many updated checklist items include multiple distinct reporting elements. As a result, assessing completeness at the item level might not indicate reliably whether all elements are reported, which creates challenges for both implementation and automated assessment. Our work complements efforts to clarify specific reporting elements and encourage more consistent and fine-grained evaluation.^10,28^

Inter-annotator agreement was reasonably high (0.782 Gwet’s AC1), suggesting that, given relevant text snippets, humans can perform the task somewhat reliably, but there might be room to improve the guidelines to promote greater reliability. Providing text snippets substantially accelerated the annotation process; however, there were a small number of cases in which the provided text did not provide sufficient context and the annotators needed to examine the full text to answer the questions.

Dataset annotation is both time-consuming and labor-intensive. A potential future direction might be to leverage reporting checklist tables that some journals require authors to submit, although their utility would depend on how accurately authors identify supporting text. Checklists that include relevant quotations or excerpts, rather than page numbers alone, may be particularly useful for reducing annotation effort and improving model performance.

Experiments in element-level assessment show that LLMs can answer fine-grained, multiple-choice questions about RCT reporting with reasonable performance in both zero-shot (F_1_: 0.822) and one-shot settings (F_1_: 0.830). The higher-capacity GPT-5 model performs slightly better than the smaller Qwen-2.5 model, though the differences are modest and confidence intervals overlap. GPT-5 achieves slightly higher sensitivity while maintaining comparable specificity, indicating that it more reliably identifies reported elements without compromising its ability to detect missing ones. These improvements come at the expense of API costs but are accompanied by faster inference (Supplementary File Section D). It is worth noting that while Qwen-2.5 underperforms GPT-5 overall, it outperforms GPT-5 on 23 of the 119 questions and achieves the same performance on 39 questions. Fine-tuning open-weight models such as Qwen-2.5 would be a promising direction in further improving model performance.

Additional analyses provided insights into factors affecting the performance and practical utility of models for LLM-based assessment. Performance was lower for questions requiring nuanced interpretation of complex or diffuse evidence (e.g., trial results assessed by CONSORT items), suggesting that future approaches may benefit from deeper contextual reasoning and evidence aggregation. Although CoT provided only modest performance gains and increased computational cost, it may still offer value by improving transparency through explicit connections between textual evidence and reporting judgments. Automatically generated question-specific prompts did not substantially improve performance, indicating that manually designed shared prompting strategies may be sufficient for many reporting assessment tasks. Detailed results from these analyses are provided in Supplementary File Section C.

As an exploratory analysis, we also assessed whether the automated approach could function as a second assessor and help reduce the burden and cost of manual assessment. The evaluation was based on six articles (three pairs) in the test set that had been independently annotated by two annotators and for which reconciled annotations were available. On this subset, the automated approach achieved a mean Gwet’s AC1 of 0.684 against the reconciled annotations. Because the model relies on automatically identified text snippets as input, whereas human annotators used expert-annotated sentences, this comparison somewhat disadvantages the model. When expert-annotated sentences were provided to the model instead, agreement increased to a mean Gwet’s AC1 of 0.763. Although still below inter-annotator agreement on this set (0.841), these results suggest that, with some refinement, the automated approach has potential to support semi-automated assessment workflows.

We formulated element-level reporting assessment as a two-step process: (1) retrieving relevant evidence snippets from the article and (2) answering a multiple-choice question based on the snippet. For evidence retrieval, our default method used the sentence-level SPIRIT-CONSORT-TM model developed in prior work.^22^ As expected, performance was lower when using automatically retrieved evidence compared with expert-annotated item-level sentence annotations (F_1_: 0.822 vs. 0.893), suggesting that enhancements to the retrieval model could further improve overall performance in element-level assessment.

To better understand these results, we also conducted experiments exploring two alternative strategies: (1) prompting the LLM to directly process the full text and (2) prompting the LLM to first identify relevant text and then use relevant text to answer each question. These experiments were conducted on a small subset (5 article pairs and 10 questions). Both approaches performed markedly worse than the default retrieval method: the full-text approach achieved an F_1_ score of 0.476, while prompting the LLM to identify relevant text first improved the F_1_ score to 0.591. As these approaches yielded lower performance while incurring substantially higher cost, we did not extend them to the full test set. These results suggest that task-specific models offer clear advantages for evidence retrieval in fine-grained RCT reporting assessment, whereas general-purpose LLMs appear less capable of performing this step reliably. This finding contrasts somewhat with recent studies suggesting that LLMs can reliably assess adherence to reporting guidelines.^17,23^ A key difference is that those studies focus on article-level adherence rather than sentence-level evidence identification. While article-level judgments may appear accurate, they are less likely to be practically useful if they are not supported by correctly identified evidence snippets, as unsupported judgments may be confusing rather than helpful for practitioners. This suggests that more attention must be paid to the identification of precise evidence supporting reporting assessments, particularly at the sentence or snippet level.

Incorporating illustrative in-context examples into the LLM prompt appears to improve performance, although performance gain depends on the example selection method and the number of in-context examples. With a single semantically similar in-context example, the model performance improves slightly (+0.8 pp F_1_), while the difference with random selection is smaller (+0.6 pp F_1_). We also observe only minor differences when using different models for calculating semantic similarity, despite having substantially different context sizes (512 tokens for SPIRIT-CONSORT-TM and 32K for *Qwen-8B-Embedding*). At the same time, using more in-context examples (3-shot and 5-shot) degraded the performance overall (results not shown). As using these specialized models for calculating semantic similarity also increases inference time significantly (especially in the case of *Qwen-8B- Embedding*), the results suggest that the utility of in-context examples might be limited for this task in practical settings.

Our qualitative error analysis revealed that LLMs generally perform well on questions that involve identifying the presence of a specific element (e.g., “Does the manuscript mention the plans for post-trial care?”). Answering such questions often amount to whether the SPIRIT-CONSORT-TM model correctly retrieves the evidence snippets, leading to an affirmative response (“Yes”). On the other hand, models struggle with questions that require deeper contextual reasoning over the retrieved relevant text snippet. For example, consider the question: “For any of the outcomes, does the manuscript report the precision around the estimated effect size?” When descriptions of results for primary and secondary outcomes are provided, LLMs sometimes treat the presence of standard errors or confidence intervals as evidence of reported precision, even when these statistics are not associated with the effect size itself. We also observed a number of error cases where conditional questions are affected by incorrect responses to the questions that they depend on. For example, ‘When reporting the results, does the manuscript mention that the adjusted analysis is pre-specified or exploratory/post hoc?’’ is dependent on an affirmative response to the question “Does the manuscript report any results from adjusted analyses?”. Consequently, an incorrect answer to the latter leads to an additional error with the former question.

Beyond predictive performance, cost, efficiency, privacy, and the need for technical expertise are important considerations when selecting an LLM for element-level assessment of RCT reports. For individual articles, API-based use was relatively inexpensive ($1.45 for GPT-5 vs. $0.45 for Qwen-2.5 via the Together AI API; https://www.together.ai) (Supplementary File Section D). At this scale, and setting aside potential privacy concerns, API access offers a practical balance of low cost, ease of use, and minimal technical overhead.

For journals, editorial teams, or meta-researchers aiming to deploy such a model at scale, per-article API costs may still remain manageable. However, alternative deployment strategies become increasingly relevant. In particular, if appropriate computational infrastructure is available, deploying open-weight models locally may be preferable. Models such as Qwen-2.5 offer performance comparable to proprietary systems while providing stronger privacy, potentially lower long-term operational costs, especially at high volume, and fine-tuning capabilities.

Our study has several limitations. First, although the corpus contains 19,500 element-level judgments, it is based on a relatively modest number of articles from RCTs with protocols and results publications in the PMC Open Access Subset, which might not be a representative sample of RCT publications more broadly. The corpus may not capture the full diversity of reporting practices across clinical specialties, journals, geographic regions, or trial designs, which may limit the portability of our findings to other settings. A more comprehensive evaluation across a broader range of articles is left for future work. Second, the best-performing model achieved an overall F_1_ score of 0.822. Although this performance is encouraging, higher performance may be necessary for reliable real-world adoption. Relatedly, while we report 95% CIs throughout, these reflect internal sampling variability within our dataset. Because our articles do not constitute a random sample from a well-defined population, the true uncertainty around our performance estimates is likely higher than the reported intervals suggest. Additional sources of uncertainty (including variability in annotator interpretation, non-deterministic LLM behavior, and corpus-specific characteristics) are not fully captured by bootstrap resampling. Third, the computational cost might be substantial in practice, as 119 questions must be answered for a single article. Although we find that an open-weight model (Qwen-2.5) achieves performance comparable to the proprietary model (GPT-5), it still requires significant GPU resources and relatively long inference times with limited computational resources. Finally, the current framework is based on the SPIRIT 2013 and CONSORT 2010 reporting guidelines. While our framework covers most items in the 2025 updated versions of these guidelines, a small number of additional questions would need to be incorporated to fully align with the updated guidelines.

Future work will focus on improving the accuracy and efficiency of the model further and developing methods to aggregate element-level outputs into interpretable summaries, making the model more practical for real-world deployment. Finally, to ensure that the framework remains aligned with evolving SPIRIT and CONSORT guidelines, we will explore automatically generating updated questions and corresponding options based on the updated reporting guidelines.

In summary, decomposing checklist items into their constituent elements more faithfully operationalizes the intent of reporting guidelines and enables a more granular characterization of RCT reporting. For authors, this granularity can produce more actionable feedback by identifying which specific elements are missing. For reviewers and journal editors, it can support more precise adherence screening by pinpointing incomplete RCT details. In addition, the framework provides higher-resolution data for meta-research, enabling systematic analysis of reporting practices and RCT characteristics across studies.

## Methods

In this section, we first describe the construction of the element-level reporting completeness framework and the SPIRIT-CONSORT-ELM dataset. We then present the automated LLM approach for assessing reporting completeness, followed by the discussion of our evaluation methodology.

### Construction of SPIRIT-CONSORT-ELM

Three experts in epidemiology and meta-research (XY, AWB, and EM-W) developed 119 questions in SPIRIT-CONSORT-ELM. These experts previously contributed to the SPIRIT-CONSORT-TM annotations and have extensive experience evaluating reporting completeness. The questions were derived from the 83 fine-grained checklist items established in SPIRIT- CONSORT-TM^22^, which in turn were based on the SPIRIT 2013^7,25^ and CONSORT 2010^1,9^ guidelines. As in SPIRIT-CONSORT-TM, we excluded some checklist items (e.g., background and interpretation) because their assessment tends to be broad and inherently subjective. To ensure consistent interpretation and annotation, the experts also developed question-specific instructions. Importantly, the framework does not assume that every reporting element applies to every trial. Rather, the questions are designed to check whether relevant considerations are discussed when applicable.

The framework includes single-choice (“check-one”), multiple-choice (“check-all-that-apply”), and multi-element questions. Multi-element questions may require either single-choice or multiple-choice responses for each constituent element. Some questions are conditionally dependent on responses to preceding questions and are presented only when applicable. These conditional questions typically request additional details when an element is present or ask for a rationale when an expected element is absent. The framework contains questions specific to protocols, results publications, or both. Examples of the different question types and applicability categories are provided in Table 4.

**Table 4.**
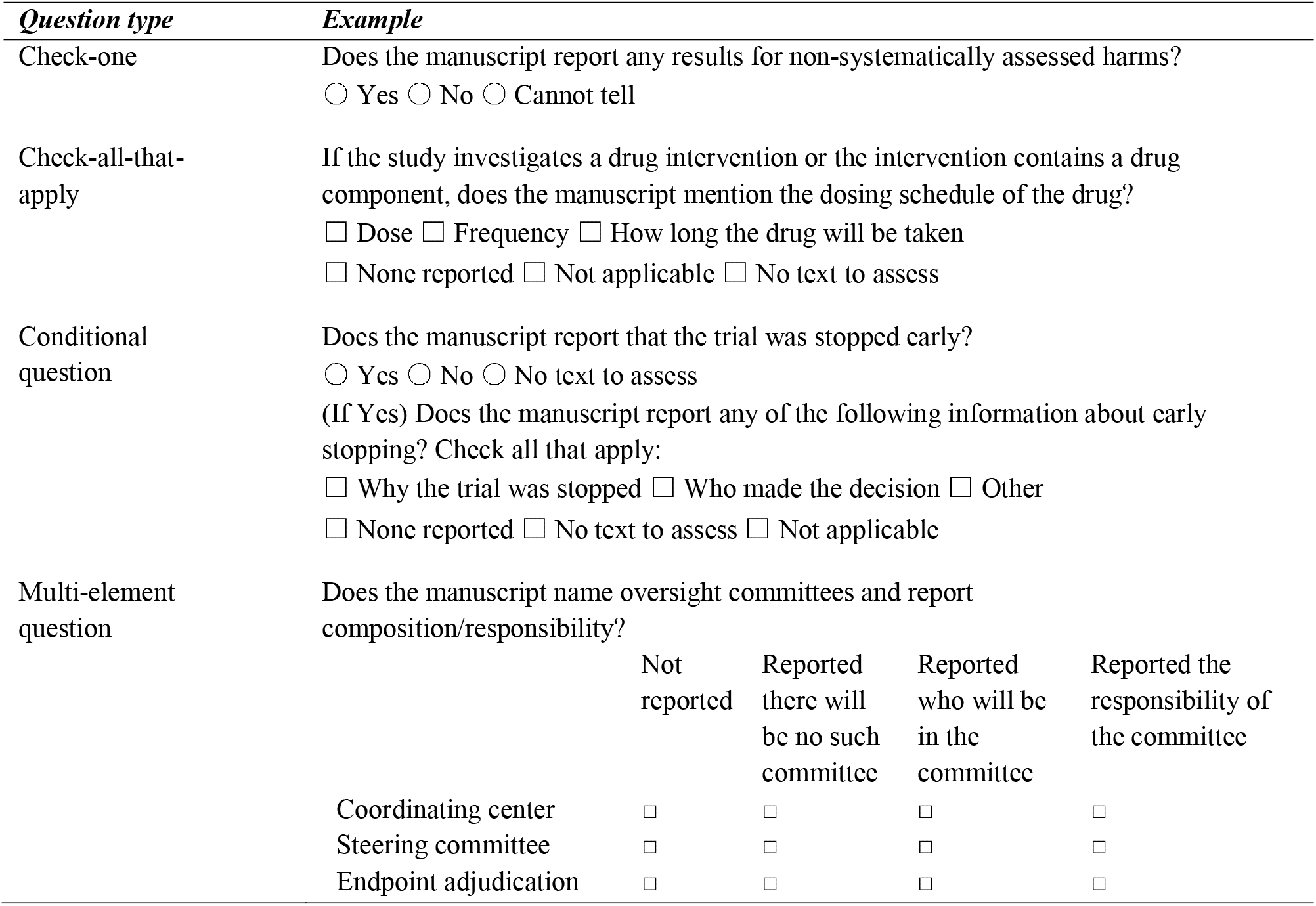
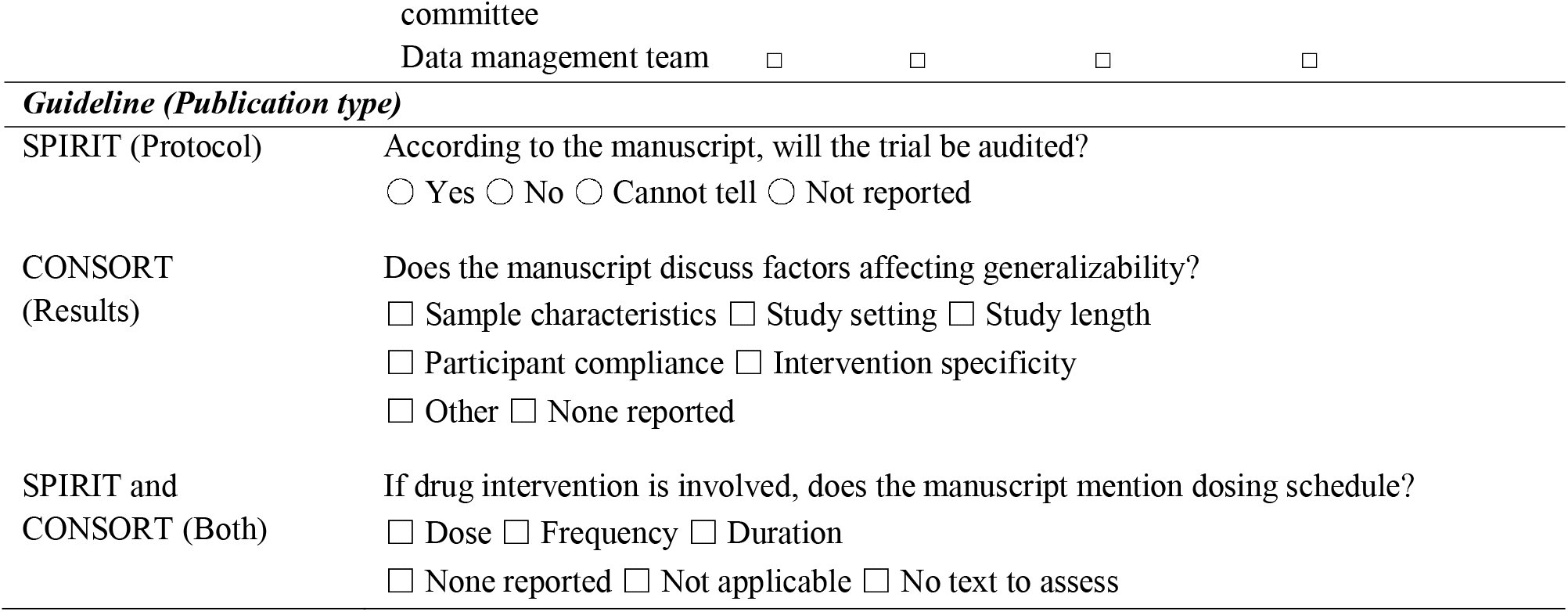
Examples of questions in SPIRIT-CONSORT-ELM. Circles indicate a single answer will be marked, while squares indicate that multiple answers can be selected. The multi-element question example requires single-choice responses for each element.

Two experts (XY and AWB) were provided with relevant text snippets for each question and completed the annotations based on these snippets. For each question, the relevant snippet comprised sentences previously annotated for the corresponding checklist item in SPIRIT- CONSORT-TM.^22^ An example annotation illustrating the relevant article text, question, question-specific instructions, and answer options is shown in Box A below. In this example, a question derived from CONSORT item 24 (Protocol) is accompanied by a sentence labeled with that checklist item. Providing relevant text snippets ensured that annotators considered only pertinent content and reduced annotation effort. All annotations were conducted using REDCap. All questions and response options are provided in Supplementary File Section B.

#### Box A. SPIRIT-CONSORT-ELM annotation example

**Relevant text:** “…As reported in the study protocol [5], we initially planned to include more than 200 participants, but we managed to randomise 56 patients only…A detailed description of the study methods has already been published [5]…. ”

**Question:** Does the manuscript mention where the protocol can be accessed (e.g., URL in the text, reference to a previous publication, article appendix, request to authors)?

##### Question-specific instruction

Answer "No" if the previous publication is a paper that describes the development of the intervention, the trial feasibility results (e.g., recruitment process), baseline characteristics, or other results.

### Options: Yes No

Importantly, annotators provided a response for a question only if the SPIRIT-CONSORT-TM dataset contained expert-annotated text snippets for the corresponding checklist item. When no relevant text was annotated or the triggering condition for a conditional question was not met, responses were automatically generated as “No”, “Not reported”, “Not applicable” or “No text to assess”, as appropriate. Specifically, we assigned “No” or “Not reported” when the question assessed reporting elements that are expected to apply to all RCTs. Otherwise, we assigned “Not applicable” when the element was determined to be not applicable based on the response to the preceding question, and “No text to assess” when applicability could not be determined. Using the example above, in the absence of any relevant text on protocol, the answer was coded as “No”.

### Inter-annotator agreement

50 articles (25 protocol-results pairs) were double-annotated by XY and AWB to assess inter-annotator agreement. The remaining 150 articles were annotated by a single annotator (XY). Inter-annotator agreement was calculated using Gwet’s AC1^29^, which adjusts for chance agreement and is preferred to Cohen’s kappa (_κ_) because it remains more stable under label imbalance. To enable these calculations, all questions were converted to a single-choice format. Specifically, each “check-all-that-apply” question was decomposed into a set of independent yes/no sub-questions, with each response treated as a separate question. For multi-element questions, each element was similarly treated as an independent question. Agreement for a given question was then computed as the proportion of options for which annotators made the same decision (yes or no). Gwet’s AC1 was then obtained by averaging across all 119 questions to avoid performance inflation by treating a single multiple-choice question as multiple separate sub-questions.

### Element-level reporting assessment using LLMs

We formulated element-level reporting assessment as a machine reading comprehension task^30^ in which a model answers structured questions based on evidence drawn from RCT publications. Our approach includes three core components: (1) evidence retrieval (identification of question-relevant text snippets); (2) machine reading comprehension using prompt engineering; and (3) use of in-context examples for question answering, selected from the training set via sentence embedding similarity. Similar to human annotation, we automatically generate responses for questions when no relevant text snippets are retrieved or the conditions for conditional questions are not met. We describe each component in more detail below and illustrate the overall pipeline in Figure 2. **The machine reading comprehension pipeline for element-level reporting assessment.** (1) evidence retrieval, (2) prompt engineering, (3) incorporating in-context examples.

**Figure 2.**
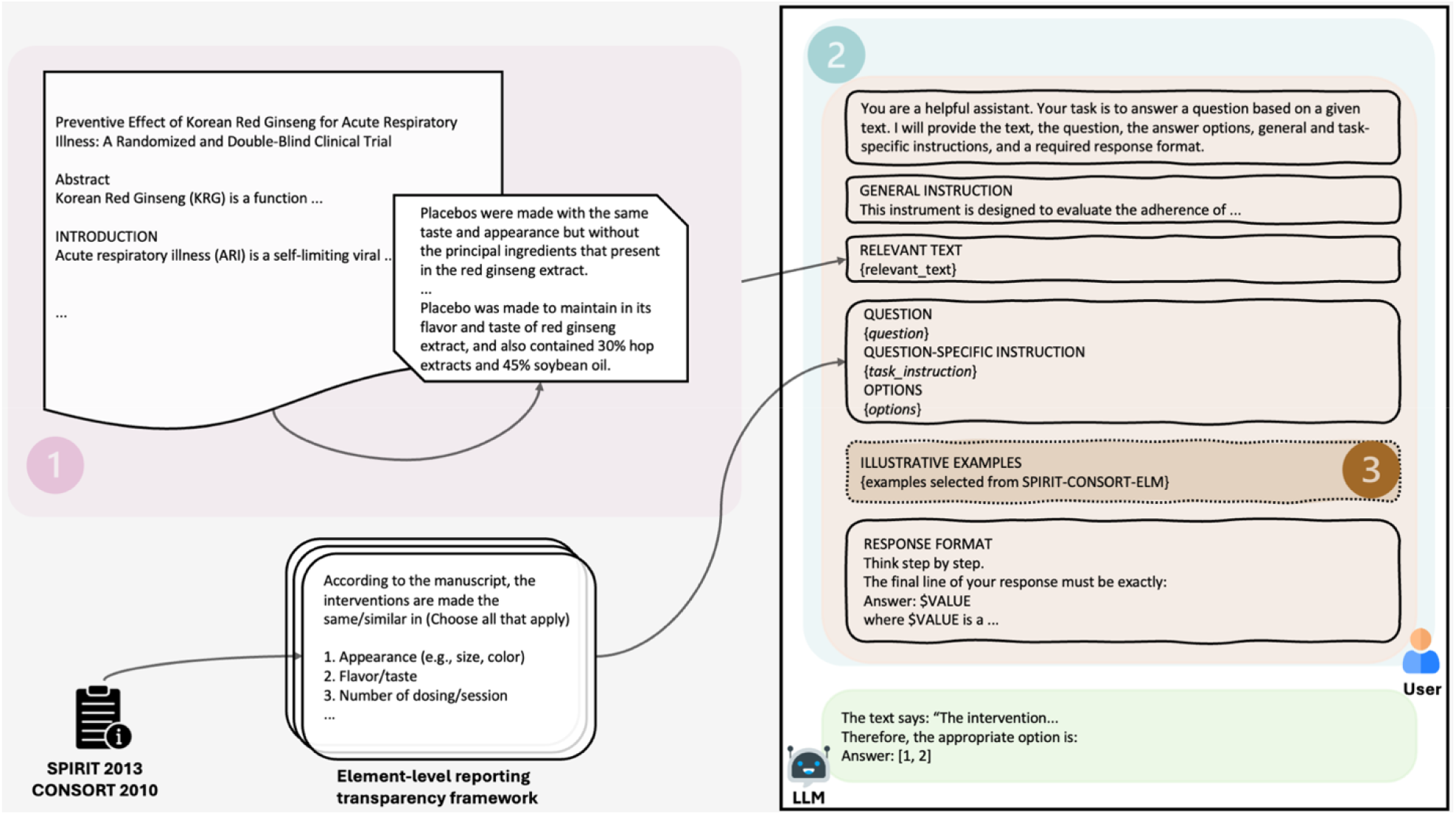
The machine reading comprehension pipeline for element-level reporting assessment. (1) evidence retrieval, (2) prompt engineering, (3) incorporating in-context examples.

#### Evidence retrieval

As the evidence source for question answering, we identify text snippets relevant to each checklist item. The primary method uses item-relevant sentences identified by the PubMedBERT model fine-tuned on the SPIRIT-CONSORT-TM dataset.^22^ This model yields a sentence-level F_1_ score of 0.74 on the test set portion of SPIRIT-CONSORT-TM.^22^ We refer to this model as SPIRIT-CONSORT-TM (in small caps). To assess the impact of evidence retrieval and to isolate the model’s predictions from errors introduced during evidence retrieval, we compare this setting to one that uses expert-annotated relevant text for each checklist item from the SPIRIT-CONSORT-TM corpus, which serves as an upper bound.

#### Machine reading comprehension using prompt engineering

Rather than construct separate LLM prompts for each question, we hand-crafted a question-agnostic prompt template that we applied uniformly across all 119 questions. The template incorporates: (1) general task description and instruction that applies to all questions, (2) relevant text snippets, (3) the question text, (4) expert-authored question-specific instructions, (5) predefined answer options, and (6) response format description (Figure 2, orange region; prompt template in Box B below). We instructed the model to select from the provided answer options only and constrained the outputs to structured responses.

We evaluated two prompting strategies. First, as shown in Box B, we incorporated a zero-shot chain-of-thought reasoning (CoT) approach by instructing the model to “Think step by step”, as prior studies have shown that encouraging intermediate reasoning can improve performance on complex tasks.^31^ As an alternative, inspired by the planning abilities of LLMs and the effectiveness of automatically generated instructions,^32,33^ in an ablation study, we also prompted the LLM to automatically generate a prompt for each question instead of using the hand-crafted prompt template. The prompt template for generating instructions is shown in Box C. An example LLM-generated instruction is provided in Supplementary File Section A.

#### Box B. Prompt template used for element-level assessment

You are a helpful assistant. Your task is to answer a question based on a given text. I will provide the text, the question, the answer options, general and task-specific instructions, and a required response format.

##### GENERAL INSTRUCTION

- This instrument is designed to evaluate the adherence of trial protocol and result papers to the SPIRIT and CONSORT reporting guidelines. It contains factual questions about the presence or absence of specific information in these documents.
- When answering the questions, rely solely on the provided texts.
- Do not include any numbers that are not part of the OPTIONS when answering the questions.
- Ensure that the numbers and the corresponding option text are consistent and aligned.

##### RELEVANT TEXT

{text}

##### QUESTION

{question_text}

##### QUESTION-SPECIFIC INSTRUCTION

{task_instruction}

##### OPTIONS

{options}

##### RESPONSE FORMAT

Think step by step.

The final line of your response must be exactly:

###### Answer: $VALUE

where $VALUE is a {response_format}.

#### Box C. Prompt template for generating instructions for each question

You are a helpful assistant. Write an instruction on how to solve the following task based on both general and task-specific instructions, all possible options of the question, and the required response format. Be sure to include clarifications for each option, avoiding any hallucinations, and be mindful of the relationships among the options. Ensure the question is explicitly included in the instruction. Follow the specified response format exactly. You are provided a section called "RELEVANT TEXT" and a placeholder {{relevant_text}} where the actual text will be inserted. Keep this section in your generation.

##### GENERAL INSTRUCTION

- This instrument is designed to evaluate the adherence of trial protocol and result papers to the SPIRIT and CONSORT reporting guidelines. It contains factual questions about the presence or absence of specific information in these documents.
- When answering the questions, rely solely on the provided texts.
- Do not include any numbers that are not part of the OPTIONS when answering the questions.
- Ensure that the numbers and the corresponding option text are consistent and aligned.

##### RELEVANT TEXT

{text}

##### QUESTION

{question_text}

##### QUESTION-SPECIFIC INSTRUCTION

{task_ instruction}

##### OPTIONS

{options}

##### RESPONSE FORMAT

Think step by step.

The final line of your response must be exactly:

###### Answer: $VALUE

where $VALUE is a {response_format}.

Prompt development was carried out using a small validation subset comprising 10 questions and 5 randomly selected article pairs from the training split. The prompt was iteratively refined and tested using GPT-5 (*gpt-5-chat-latest*) until the best-performing configuration was identified.

#### In-context examples

To leverage the LLM’s in-context learning capabilities, we augmented the hand-crafted template with illustrative examples selected from the portion of the dataset not used for testing. We explored two example selection strategies: *random selection* and *semantic similarity–based selection*. Under the random selection strategy, for each question we sampled one training instance per answer option and included it as an illustrative example to ensure coverage of all possible options. Under the semantic similarity–based strategy, we dynamically retrieved examples whose relevant text snippets were most similar to the snippet used to answer the question, with similarity computed from text embeddings. We evaluated two types of embedding models: encoder-based (SPIRIT-CONSORT-TM model^22^), and decoder-based (*Qwen-8B-Embedding*^34^). The encoder model supports deep contextual understanding but is limited to 512 tokens. The decoder-based model is able to retrieve examples with longer context.

### Evaluation of automated approach

In this section, we describe the experimental settings, evaluation strategies, and evaluation metrics. We used two base LLMs: GPT-5 and the open-weight model Qwen-2.5 (*qwen2.5- 72b-instruct*) under identical experimental conditions. We accessed GPT-5 through API and ran an 8-bit quantized version of Qwen-2.5 using *llama.cpp*^35^ on a single NVIDIA H100 GPU. For the best performing model, we analyzed the performance broken down by question type (check-one vs. check-all-that-apply) and relevant guideline (SPIRIT vs. CONSORT). We also conducted a series of ablation studies to evaluate the impact of alternative design choices at each stage. All experiments were conducted from February 3, 2026 to February 20, 2026.

All evaluations were conducted on the test split of SPIRIT-CONSORT-TM (20 protocol– results pairs) to ensure consistency and avoid data leakage. This is necessary because the primary evidence retrieval component (the SPIRIT-CONSORT-TM model) was fine-tuned on the training split of the SPIRIT-CONSORT-TM dataset; evaluating on the training data would risk reproducing expert-annotated text and inflating performance estimates.

We used a majority class baseline for comparison with LLM performance. For each question, the most frequent option was determined from the 80 article pairs excluded from the test set and assigned to all corresponding test instances.

As primary evaluation metrics for LLM-based element-level reporting assessment, we used the F_1_ score, sensitivity (recall), specificity, and Gwet’s AC1. The F_1_ score is the harmonic mean of precision (positive predictive value; PPV) and sensitivity and ranges from 0 to 1 (the higher the better). Sensitivity measures the proportion of true positives correctly identified, whereas specificity measures the proportion of true negatives correctly identified. Gwet’s AC1 quantifies agreement between model predictions and expert annotations while accounting for chance agreement. As secondary metrics, we report per-question F_1_ score, precision, sensitivity, specificity, and negative predictive value (NPV). For multiple-answer (‘check-all-that-apply’) questions, which cannot be evaluated directly using F_1_ and Gwet’s AC1 metrics, we decomposed each question into multiple binary sub-items and computed a micro-averaged F_1_ score and averaged Gwet’s AC1. Final results are reported as mean scores with 95% confidence intervals (CIs) across all 119 questions, estimated using bootstrap resampling with 10,000 iterations.

In addition to performance metrics, we also measured the average time it takes to process an article for all 119 questions and the cost per article to assess practical feasibility.

## Data availability

The data used for all experiments and analyses in this study are publicly available at https://osf.io/kznx4/.

## Code availability

The code used for all experiments and analyses in this study are publicly available at https://osf.io/kznx4/.

## Data Availability

All data produced are available online at https://osf.io/kznx4/.

## Acknowledgements

This work was supported by the National Library of Medicine of the National Institutes of Health under the award number R01LM014079. The content is solely the responsibility of the authors and does not necessarily represent the official views of the National Institutes of Health or any other organization. The funder had no role in considering the study design or in the collection, analysis, interpretation of data, writing of the report, or decision to submit the article for publication. AWB is supported in part by the Arkansas Children’s Research Institute and the Arkansas Biosciences Institute. This work used Bridges-2 and Ocean at Pittsburgh Supercomputing Center (PSC) through allocation #CIS230380 from the Advanced Cyberinfrastructure Coordination Ecosystem: Services & Support (ACCESS) program^36^, which is supported by National Science Foundation, United States grants #2138259, #2138286, #2138307, #2137603, and #2138296.

## Author Contributions

L.J.: Methodology, Data curation, Software, Validation, Formal analysis, Investigation, Writing – Original draft, Writing – Review & Editing. X.Y.: Conceptualization, Methodology, Data curation, Writing – Review & Editing. A.W.B.: Conceptualization, Methodology, Data curation, Writing – Review & Editing. M.L.: Methodology, Software, Validation, Writing – Original draft, Writing – Review & Editing. W.S.: Software, Formal analysis. J.D.M.: Writing – Review & Editing. C.J.V: Writing – Review & Editing. E.M.- W.: Conceptualization, Methodology, Data curation, Supervision, Project administration, Funding acquisition, Writing – Review & Editing. H.K.: Conceptualization, Methodology, Investigation, Supervision, Project administration, Funding acquisition, Writing – Original draft, Writing – Review & Editing. All authors have read and approved the manuscript.

---

**Figure 3. Performance comparison of the LLM approaches and the majority class baseline**. F_1_, sensitivity, specificity, and Gwet’s AC1 scores are reported for 119 questions along with 95% CIs. LLM approaches use SPIRIT-CONSORT-TM-identified snippets and hand-crafted prompt template. CI: confidence interval.

**Figure 4. The machine reading comprehension pipeline for element-level reporting assessment.** (1) evidence retrieval, (2) prompt engineering, (3) incorporating in-context examples.

