## Supplementary Material for "SPIRIT-CONSORT-ELM: Element-Level Annotated Dataset and Large Language Model Approach for Assessing Randomized Controlled Trial Reporting"

### **Section A. LLM-generated instruction for an element-level question (“[Results] Does the manuscript report any results for non-systematically assessed harms?”)**

**INSTRUCTION**

You are asked to answer the following question based on the provided **RELEVANT TEXT**:

**QUESTION**

56a. [Results] Does the manuscript report any results for non-systematically assessed harms?

**STEP-BY-STEP GUIDANCE**

**Read the RELEVANT TEXT carefully.**

Use only the information that appears in the provided text section. Do not infer information from outside sources.

**Identify mentions of harms or adverse events.**

Look for terms such as “harms,” “adverse events,” “side effects,” or “safety outcomes.”

**Determine whether these harms were assessed non-systematically.**

Harms are **likely non-systematically assessed** when the manuscript refers to **spontaneously reported** events,

**MedDRA**, or other systems for classifying unscheduled reported data.

The results for these harms are typically presented as **number of events** or **number of people with the event** rather than as continuous measures (means or medians).

**Distinguish this from systematically assessed harms.** - Harms are **systematically assessed** if results come from **predefined times** and measurements for **all participants**, often shown as means or medians.

**Check if any results for non-systematic harms are actually reported.**

Results may be in tables or described in the text.

Even if the text refers to a figure or supplement, **only use what is explicitly described**; do not assume contents not provided in the text.

**INTERPRET THE OPTIONS CAREFULLY**

**Option 1 (Yes):**

Choose this if the text explicitly reports any results for harms that were assessed non-systematically (for example, spontaneous reports summarized with counts or numbers of affected individuals).

**Option 2 (No):**

Choose this if the text does **not** report any non-systematically assessed harms, meaning no mention of spontaneously reported harms or any results of such nature.

**Option 3 (Cannot tell):**

Choose this if it is **unclear** whether any reported harms were non-systematically assessed — for instance, if harms are mentioned but details on how they were collected or reported are insufficient to decide.

**Ensure consistency and completeness:**

Use only the information in the RELEVANT TEXT placeholder.

Do not introduce external information or speculate.

Choose **only one option (1, 2, or 3)**.

The numbers used must match exactly the given option numbers.

**RELEVANT TEXT**

{relevant_text}

**RESPONSE FORMAT**

Think step by step.

The final line of your response must be exactly:

**Answer: $VALUE**

where **$VALUE** is replaced with **1**, **2**, or **3**, corresponding to your final choice.

### **Section B. Element-level assessment**

In this section, we present the 119 questions, their associated options, and the performance of the primary LLM pipeline (Table S1). This pipeline uses the spirit-consort-tm model for evidence retrieval, a hand-crafted prompt template for machine reading comprehension, and no in-context examples.

**Table S1**. The list of 119 questions, relevant options, and per-question GPT-5 performance. PPV: positive predictive value; NPV: negative predictive value.

| **ID** | **Question** | **Options** | **F_1_** | **GWET** | **Sensitivity** | **Specificity** | **PPV** | **NPV** |
| --- | --- | --- | --- | --- | --- | --- | --- | --- |
| 1a | [Results] Does the manuscript mention where the protocol can be accessed (e.g., URL in the text, reference to a previous publication, article appendix, request to authors)? | $○$ Yes $○$ No | 0.70 | 0.52 | 0.70 | 0.70 | 0.70 | 0.70 |
| 1b | [Results] Does the manuscript mention where the statistical analysis plan can be accessed (e.g., URL in the text, reference to a previous publication, article appendix, request to authors)? | $○$ Yes $○$ No | 1.00 | 1.00 | 1.00 | 1.00 | 1.00 | 1.00 |
| 2a | According to the manuscript, does the trial receive any financial, material, or other support? | $○$ Yes $○$ No $○$ Not reported | 0.88 | 0.86 | 0.88 | 0.94 | 0.88 | 0.94 |
| 2b | [If 2a=Yes] Does the manuscript mention whether the funder is involved in the following? \|; Study design \|; Data collection, management, analysis, or interpretation \|; Writing of the report \|; Decision to submit the report for publication \| | Not reported \| Reported “involved” \| Reported “not involved”; $○$ \| $○$ \| $○$; $○$ \| $○$ \| $○$; $○$ \| $○$ \| $○$; $○$ \| $○$ \| $○$ | 0.93 | 0.90 | 0.93 | 0.94 | 0.93 | 0.94 |
| 2c | [If 2a=Yes] Does the manuscript mention any non-financial (e.g., equipment, drugs, services) support? | $○$ Yes $○$ No $○$ Not reported $○$ No text to assess | 0.78 | 0.74 | 0.93 | 0.94 | 0.93 | 0.94 |
| 3a | Does the manuscript name the sponsor? | $○$ Yes $○$ No $○$ Cannot tell | 0.93 | 0.91 | 0.93 | 0.94 | 0.93 | 0.94 |
| 3b | [If 3a=Yes] Does the manuscript mention the role of the sponsor in the study (e.g., involvement in the study design, data collection, management, analysis, and interpretation, writing of the report, decision to submit the report for publication, etc.)? | $○$ Yes $○$ No | 1.00 | 1.00 | 0.93 | 0.94 | 0.93 | 0.94 |
| 4a | Does the manuscript mention authors’ contribution to one or more of the following? Check all that apply. | □ Conceptualization □ Investigation □ Data curation □ Formal analysis □ Writing - Original Draft □ Writing - Review & Editing □ Supervision □ Project administration □ Funding acquisition □ Other □ None reported | 0.94 | 0.92 | 0.78 | 0.93 | 0.78 | 0.93 |
| 5a | Does the manuscript name one or more of the following oversight committees? And if yes, does the manuscript mention its composition and responsibility? \|; Coordinating center \|; Steering committee \|; Endpoint adjudication committee \|; Data management team \| | Not reported \| Reported there will be no such committee \| \| Reported who will be in the committee \| Reported the responsibility of the committee; □ \| \| □ \| □ \| □; □ \| \| □ \| □ \| □; □ \| \| □ \| □ \| □; □ \| \| □ \| □ \| □ | 0.94 | 0.96 | 0.93 | 0.93 | 0.93 | 0.93 |
| 5b | Does the manuscript mention any other individuals or groups who will oversee the trial? | $○$ Yes $○$ No $○$ Cannot tell | 0.88 | 0.86 | 1.00 |  | 1.00 |  |
| 6a | Does the manuscript include the following information in the study objective/hypothesis? Check all that apply. | □ Intervention □ Comparator □ Outcome □ Population □ None reported | 0.89 | 0.79 | 0.96 | 0.94 | 0.90 | 0.98 |
| 7a | Does the manuscript mention the study setting? Check all that apply. | □ Healthcare setting □ Community-based □ Academic or research institutions □ Geographical characteristics □ Social, economic, and cultural environment □ Other □ Can’t tell □ None reported | 0.71 | 0.79 | 0.82 | 0.88 | 0.68 | 0.89 |
| 7b | Does the manuscript mention the study’s geographical location, or if applicable, provide a reference to where a list of the names of participating sites/countries can be found? | $○$ Yes $○$ No | 0.80 | 0.74 | 0.80 | 0.80 | 0.80 | 0.80 |
| 8a | Does the manuscript mention the following in the participant eligibility criteria? Check all that apply. | □ Disease criteria that specify clinical parameters of the disease being studied □ Precision criteria that render the study population more homogeneous for the purposes of the trial □ Safety criteria that exclude persons thought to be unduly vulnerable to harm from the study therapy □ Administrative criteria that ensure the smooth functioning of the trial □ None reported | 0.77 | 0.59 | 0.75 | 0.78 | 0.78 | 0.64 |
| 9a | Does the manuscript mention the eligibility criteria for study centers/sites to participate in the study? | $○$ Yes $○$ No | 0.93 | 0.92 | 0.93 | 0.93 | 0.93 | 0.93 |
| 9b | Does the manuscript mention the eligibility criteria for interventionists to participate in the study? | $○$ Yes $○$ No | 0.93 | 0.92 | 0.93 | 0.93 | 0.93 | 0.93 |
| 10a | If the study investigates a drug intervention or the intervention contains a drug component, does the manuscript mention the dosing schedule of the drug? Check all that apply. | □ Dose □ Frequency □ How long the drug will be taken □ None reported □ Not applicable □ No text to assess | 0.85 | 0.88 | 0.82 | 0.97 | 0.88 | 0.93 |
| 10b | If the study investigates a non-drug intervention or the intervention contains a non-drug component, does the manuscript mention the following of the intervention? Check all that apply. | □ Physical or informational materials used in the intervention □ Procedures, activities, and/or processes used in the intervention □ Any specific training given to the intervention providers □ Modes of delivery □ Number of times the intervention delivered, including the number of sessions, their schedule, and their duration or intensity □ Intervention delivered over what time period □ Other □ None reported □ Not applicable □ No text to assess | 0.83 | 0.80 | 0.88 | 0.87 | 0.80 | 0.93 |
| 10c | If the intervention or control contains usual care or standard of care, does the manuscript describe what constitutes ’usual care’ or ’standard of care’? | $○$ Yes $○$ No $○$ Not applicable $○$ No text to assess | 0.85 | 0.83 | 0.85 | 0.95 | 0.85 | 0.95 |
| 11a | Does the manuscript mention any of the following reasons for modifying or discontinuing the allocated intervention, or for withdrawing participants from the study? Check all that apply. | □ In response to harms □ Participant request □ Improving/worsening disease □ At investigator’s discretion □ Other □ None reported | 0.73 | 0.89 | 0.98 | 0.87 | 0.58 | 0.91 |
| 12a | Does the manuscript mention the procedures for monitoring participant adherence to the intervention? | $○$ Yes $○$ No $○$ Cannot tell | 0.93 | 0.86 | 0.93 | 0.93 | 0.93 | 0.93 |
| 12b | Does the manuscript mention any strategies (other than monitoring adherence) to enhance participant adherence to the intervention? | $○$ Yes $○$ No $○$ Cannot tell | 0.88 | 0.85 | 0.88 | 0.88 | 0.88 | 0.88 |
| 12c | [Results] Does the manuscript report results concerning participant adherence to the intervention? | $○$ Yes $○$ No $○$ Not applicable $○$ Cannot tell $○$ No text to assess | 0.70 | 0.56 | 0.70 | 0.85 | 0.70 | 0.85 |
| 12d | For non-drug interventions, does the manuscript mention the procedures for monitoring interventionist’s adherence to the intervention? | $○$ Yes $○$ No $○$ Not applicable $○$ Cannot tell $○$ No text to assess | 0.62 | 0.52 | 0.62 | 0.88 | 0.62 | 0.88 |
| 12e | For non-drug interventions, does the manuscript mention any strategies (other than monitoring adherence) to improve interventionist’s adherence to the intervention (e.g., interventionist training)? | $○$ Yes $○$ No $○$ Not applicable $○$ Cannot tell $○$ No text to assess | 0.53 | 0.39 | 0.53 | 0.84 | 0.53 | 0.84 |
| 12f | [Results] For non-drug interventions, does the manuscript report results concerning interventionist’s adherence to the intervention? | $○$ Yes $○$ No $○$ Not applicable $○$ Cannot tell $○$ No text to assess | 0.65 | 0.56 | 0.65 | 0.88 | 0.65 | 0.88 |
| 13a | Does the manuscript mention the concomitant care and interventions for only one group or some of the groups while omitting this information for the other group(s)? | $○$ Yes $○$ No $○$ Cannot tell $○$ Not reported | 0.72 | 0.64 | 0.72 | 0.86 | 0.72 | 0.86 |
| 14a | [Protocol] Does the manuscript mention the plans for ancillary care during the trial? | $○$ Yes $○$ No $○$ Cannot tell | 0.95 | 0.95 | 0.95 | 0.95 | 0.95 | 0.95 |
| 14b | [Protocol] Does the manuscript mention the plans for post-trial care? | $○$ Yes $○$ No $○$ Cannot tell | 0.95 | 0.95 | 0.95 | 0.95 | 0.95 | 0.95 |
| 14c | [Protocol] Does the manuscript mention the plans to compensate participants for trial-related harms? | $○$ Yes $○$ No $○$ Cannot tell | 0.80 | 0.76 | 0.80 | 0.80 | 0.80 | 0.80 |
| 14d | [Protocol] If "No" to at least one of 14a,14b, and 14c, does the manuscript mention a rationale to justify why there is no provision for ancillary care, post-trial care, or compensation for trial-related harms? | $○$ Yes $○$ No $○$ Cannot tell $○$ No text to assess $○$ Not applicable | 0.75 | 0.68 | 0.75 | 0.75 | 0.75 | 0.75 |
| 15a | Does the manuscript specify one or more outcomes as the primary outcome? | $○$ Yes $○$ No | 0.80 | 0.68 | 0.80 | 0.80 | 0.80 | 0.80 |
| 15b | [If 15a=Yes] For the primary outcome or the first-mentioned primary outcome if there is more than one primary outcome, does the manuscript report the following for that outcome? Check all that apply. | □ Specific measurement □ Analysis metric □ Method of aggregation □ Measurement time point of interest □ None reported □ No text to assess | 0.75 | 0.72 | 0.74 | 0.85 | 0.68 | 0.86 |
| 15c | [If 15a=Yes] For the primary outcome or the first-mentioned primary outcome if there is more than one primary outcome, does the manuscript mention the rationale of choosing that outcome? | $○$ Yes $○$ No $○$ No text to assess | 0.65 | 0.48 | 0.65 | 0.82 | 0.65 | 0.82 |
| 15d | Does the manuscript specify one or more outcomes as the non-primary outcome? | $○$ Yes $○$ No | 0.80 | 0.71 | 0.80 | 0.80 | 0.80 | 0.80 |
| 15e | [If 15d=Yes] For the first-mentioned non-primary outcomes, does the manuscript report the following for that outcome? Check all that apply. | □ Specific measurement □ Analysis metric □ Method of aggregation □ Measurement time point of interest □ None reported □ No text to assess | 0.56 | 0.63 | 0.69 | 0.70 | 0.42 | 0.95 |
| 15f | [If 15d=Yes] For the first-mentioned non-primary outcomes, does the manuscript mention the rationale of choosing that outcome? | $○$ Yes $○$ No $○$ No text to assess | 0.53 | 0.38 | 0.53 | 0.76 | 0.53 | 0.76 |
| 16a | [Results] Does the manuscript mention any changes to trial outcomes after the trial has started? | $○$ Yes $○$ No $○$ Cannot tell $○$ No text to assess | 0.90 | 0.89 | 0.90 | 0.90 | 0.90 | 0.90 |
| 16b | [Results, if 16a=Yes] What changes are reported? Check all that apply. | □ Change in the way that an outcome is assessed □ Original outcome replaced with new outcome □ Original outcome dropped □ New outcome added □ Change in the designation of outcomes as primary or secondary □ Other □ Can’t tell □ No text to assess □ Not applicable | 0.90 | 0.97 | 1.00 | 1.00 | 0.90 | 0.99 |
| 16c | [Results, if 16a=Yes] Does the manuscript mention the rationale for one or more of the changes? | $○$ Yes $○$ No $○$ Cannot tell $○$ No text to assess $○$ Not applicable | 0.90 | 0.89 | 0.90 | 0.90 | 0.90 | 0.90 |
| 17a | Given the available information from texts and diagrams, if either or both are present, does the participant timeline meet the following criteria? Check all that apply. | □ Start from initial eligibility screening through to study close-out □ Include timing of each visit □ Include time points or periods during which trial interventions will be administered □ Include the procedures performed at each visit □ Include the assessments performed at each visit □ Other □ None reported □ Supplementary figures/tables | 0.57 | 0.67 | 0.53 | 0.86 | 0.55 | 0.85 |
| 17b | Is the timeline depicted in a schematic diagram (e.g., figure, table)? | $○$ Yes $○$ No $○$ Cannot tell | 0.78 | 0.55 | 0.78 | 0.78 | 0.78 | 0.78 |
| 18a | Does the manuscript describe a formal sample size calculation process for determining or partially determining the sample size? | $○$ Yes $○$ No $○$ Cannot tell $○$ Not reported | 0.97 | 0.97 | 0.97 | 0.99 | 0.97 | 0.99 |
| 18b | [If 18a=Yes] Is any of the following specified? Check all that apply. | □ The outcome on which the calculation was based □ Effect size □ Justification or reference of the effect size □ Type I error or confidence interval level □ One-sided or two-sided □ Type II error or power □ Statistical test □ Any allowance made for attrition or non-compliance during the study □ Other □ None reported | 0.89 | 0.81 | 0.93 | 0.83 | 0.77 | 0.96 |
| 19a | Does the manuscript mention the following information of the recruitment process? Check all that apply. | □ Where participants will be recruited □ When participants will be recruited □ How participants will be recruited □ Expected recruitment rates or duration of the recruitment period □ Plans to monitor recruitment during the trial □ Any financial or non-financial incentives provided to trial investigators or participants for enrolment □ Other □ None reported | 0.81 | 0.87 | 0.89 | 0.94 | 0.89 | 0.94 |
| 20a | Once potential participants express interest in the study, they undergo an eligibility assessment. Does the manuscript mention who will assess a potential participant’s eligibility? | $○$ Yes $○$ No $○$ Cannot tell | 0.80 | 0.77 | 0.80 | 0.90 | 0.80 | 0.90 |
| 20b | After participants have been recruited and assessed for eligibility, those who meet the criteria and consent to participate are formally enrolled in the study. Does the manuscript mention who will enroll participants to the study? | $○$ Yes $○$ No $○$ Cannot tell | 0.72 | 0.68 | 0.72 | 0.86 | 0.72 | 0.86 |
| 21a | Does the manuscript mention the following detail about the consent process? Check all that apply. | □ Who obtains the consent □ Status/experience/training of the research team members who obtain the consent □ How pertinent information is provided to potential participants □ How potential participant’s understanding of the materials is assessed □ Explanation as to why the consent process from the participant is modified or waived □ Other □ None reported | 0.52 | 0.70 | 0.72 | 0.88 | 0.67 | 0.87 |
| 22a | Does the manuscript provide a model consent form as an appendix? | $○$ Yes $○$ No $○$ Cannot tell | 1.00 | 1.00 | 1.00 |  | 1.00 |  |
| 23a | [Protocol] According to the manuscript, will there be additional consent provisions for collection and use of participant data and biological specimens in ancillary studies? | $○$ Yes $○$ No $○$ Cannot tell $○$ No text to assess | 1.00 | 1.00 | 1.00 |  | 1.00 |  |
| 23b | [Protocol, if 23a=Yes] Does the manuscript mention further details about the additional consent processes? | $○$ Yes $○$ No $○$ No text to assess $○$ Not applicable | 1.00 | 1.00 | 1.00 |  | 1.00 |  |
| 24a | According to the manuscript, is the allocation sequence generated (check all that apply) | □ By a person □ By a centralized website/system □ At a specified location □ Other □ None reported | 0.65 | 0.76 | 0.89 | 0.94 | 0.89 | 0.94 |
| 24b | Does the manuscript mention that the random sequence generation process is separate from the rest of the research team? | $○$ Yes $○$ No $○$ Cannot tell | 0.78 | 0.69 | 0.64 | 0.89 | 0.60 | 0.91 |
| 25a | Does the manuscript mention conditions (e.g., serious adverse events that require immediate medical intervention) under which emergency unmasking can occur? | $○$ Yes $○$ No $○$ Cannot tell | 0.93 | 0.92 | 0.78 | 0.89 | 0.78 | 0.89 |
| 25b | Does the manuscript mention the procedures for revealing a participant’s allocated intervention? | $○$ Yes $○$ No $○$ Cannot tell | 0.95 | 0.95 | 0.93 | 0.93 | 0.93 | 0.93 |
| 26a | According to the manuscript, the interventions are made the same/similar in (Choose all that apply) | □ Appearance □ Flavor/taste □ Number of dosing/session □ Timing of dosing/session □ Duration of the intervention □ Other aspects □ None reported □ Can’t tell | 0.88 | 0.96 | 0.95 | 0.95 | 0.95 | 0.95 |
| 27a | Does the manuscript mention who will collect any of the outcome, baseline, or other trial data? | $○$ Yes $○$ No $○$ Cannot tell | 0.62 | 0.26 | 0.79 | 0.94 | 0.97 | 0.98 |
| 27b | Does the manuscript mention the methods used to collect any of the outcome, baseline, or other trial data? | $○$ Yes $○$ No $○$ Cannot tell | 0.80 | 0.67 | 0.62 | 0.62 | 0.62 | 0.62 |
| 27c | Does the manuscript mention any of the following strategies to improve data quality during the data collection process? Check all that apply. | □ Training/certification of data collection personnel □ Masking of data collection personnel □ Use of standardized methods or instruments with high validity or reliability □ Duplicate data measurements □ Pilot testing data collection forms □ Central lab testing □ Other □ None reported | 0.63 | 0.80 | 0.80 | 0.80 | 0.80 | 0.80 |
| 28a | Does the manuscript mention any of the following strategies to promote participant retention? Check all that apply. | □ Financial reimbursement □ Multiple contact attempts □ Send out reminders □ Monitor retention □ Multiple follow-up approaches □ Flexible scheduling options □ Other □ None reported | 0.86 | 0.95 | 0.53 | 0.89 | 0.58 | 0.88 |
| 28b | Does the manuscript mention whether outcome data for non-adherence and non-retention will still be collected? | $○$ Yes $○$ No $○$ Cannot tell | 0.90 | 0.89 | 0.74 | 0.96 | 0.97 | 0.98 |
| 29a | Does the manuscript mention the following aspect of data management process? Check all that apply. | □ Data entry □ Data coding process □ Data security □ Data storage □ None reported | 0.95 | 0.97 | 0.90 | 0.95 | 0.90 | 0.95 |
| 29b | Does the manuscript mention any strategies to promote data quality during the data management processes? Check all that apply. | □ Double data entry □ Data checking □ Other □ None reported | 0.98 | 0.99 | 0.94 | 0.98 | 0.93 | 0.99 |
| 30a | Does the manuscript mention the statistical methods used explicitly for comparing the primary outcome (or any of the primary outcomes if there are more than one)? | $○$ Yes $○$ No $○$ Cannot tell | 0.65 | 0.52 | 1.00 | 0.99 | 0.92 | 1.00 |
| 30b | Does the manuscript identify the primary analysis for the study? | $○$ Yes $○$ No $○$ Not applicable $○$ Cannot tell $○$ No text to assess | 0.65 | 0.59 | 0.65 | 0.88 | 0.65 | 0.88 |
| 31a | Does the manuscript mention that subgroup analysis will be/was conducted? | $○$ Yes $○$ No | 0.82 | 0.73 | 0.82 | 0.82 | 0.82 | 0.82 |
| 31b | [Protocol, if 31a=Yes] Is subgroup analysis specified with further details? | $○$ Yes $○$ No $○$ Cannot tell $○$ No text to assess | 0.80 | 0.75 | 0.80 | 0.90 | 0.80 | 0.90 |
| 31c | [Results, if 31a=Yes] Does the manuscript mention whether the subgroup analysis is pre-specified or exploratory/post hoc? | $○$ Yes $○$ No $○$ Cannot tell $○$ No text to assess | 0.80 | 0.77 | 0.80 | 0.93 | 0.80 | 0.93 |
| 31d | Does the manuscript mention that adjusted analysis will be/was conducted? | $○$ Yes $○$ No | 0.78 | 0.63 | 0.78 | 0.78 | 0.78 | 0.78 |
| 31e | [Protocol, if 31d=Yes] Is adjusted analysis specified with further details? | $○$ Yes $○$ No $○$ Cannot tell $○$ No text to assess | 0.75 | 0.70 | 0.75 | 0.88 | 0.75 | 0.88 |
| 31f | [Results, if 31d=Yes] Does the manuscript mention whether the adjusted analysis is pre-specified or exploratory/post hoc? | $○$ Yes $○$ No $○$ Cannot tell $○$ No text to assess | 0.60 | 0.52 | 0.60 | 0.87 | 0.60 | 0.87 |
| 32a | According to the manuscript, does the trial have a data monitoring committee? | $○$ Yes $○$ No $○$ Cannot tell $○$ Not reported | 0.95 | 0.94 | 0.95 | 0.97 | 0.95 | 0.97 |
| 32b | [If 32a=No] Does the manuscript mention the rationale for not having a data monitoring committee? | $○$ Yes $○$ No $○$ Not applicable | 0.97 | 0.96 | 0.97 | 0.97 | 0.97 | 0.97 |
| 32c | [If 32a=Yes] Does the manuscript mention the following aspects of the data monitoring committee? Check all that apply. | □ Composition or intended size and characteristics of the membership □ Roles and responsibilities □ Planned method of functioning □ Degree of independence from the sponsor and investigators □ Other □ None reported □ No text to assess □ Not applicable | 0.88 | 0.95 | 0.72 | 0.99 | 0.86 | 0.96 |
| 33a | According to the manuscript, does the trial perform interim analyses? | $○$ Yes $○$ No $○$ Cannot tell $○$ Not reported | 0.93 | 0.91 | 0.93 | 0.96 | 0.93 | 0.96 |
| 33b | [If 33a=Yes] Does the manuscript mention the following aspects of the interim analyses? Check all that apply. | □ Rationale for the analysis □ Statistical methods used for the analysis □ Timing of the analysis □ Who will conduct the analysis □ If the analysis will be masked □ Who will have the access to the analysis results □ Whether those with access to the results will remain masked □ Whether masking will be maintained when any adaptations to the trial are made □ Other □ None reported □ No text to assess □ Not applicable | 0.88 | 0.97 | 0.76 | 0.97 | 0.99 | 0.98 |
| 34a | According to the manuscript, does the trial have a stopping rule? | $○$ Yes $○$ No $○$ Cannot tell $○$ Not reported | 0.90 | 0.89 | 0.90 | 0.95 | 0.90 | 0.95 |
| 34b | [If 34a=Yes] Does the manuscript mention who has the ultimate authority to stop the trial? | $○$ Yes $○$ No $○$ No text to assess $○$ Not applicable | 0.85 | 0.84 | 0.85 | 0.95 | 0.85 | 0.95 |
| 35a | Does the manuscript mention that non-systematically assessed harms will be/was collected? | $○$ Yes $○$ No $○$ Cannot tell | 0.82 | 0.79 | 0.82 | 0.91 | 0.82 | 0.91 |
| 35b | Does the manuscript mention how the collected data of non-systematically assessed harms will be/was analyzed and reported in the result reports? | $○$ Yes $○$ No $○$ Cannot tell | 1.00 | 1.00 | 1.00 |  | 1.00 |  |
| 36a | [Protocol] According to the manuscript, will the trial be audited? | $○$ Yes $○$ No $○$ Cannot tell $○$ Not reported | 0.95 | 0.95 | 0.95 | 0.97 | 0.95 | 0.97 |
| 36b | [Protocol, if 36a=Yes] Does the manuscript mention the following aspects of trial auditing? Check all that apply. | □ Procedures of auditing □ Frequency of auditing □ Personnel involved and their degree of independence from the trial investigators and sponsor □ Other □ None reported □ No text to assess □ Not applicable | 0.93 | 0.98 | 1.00 | 0.93 | 0.98 | 0.99 |
| 37a | Does the manuscript mention where ethics approval has been obtained, or plans to seek such approval? | $○$ Yes $○$ No $○$ Not applicable $○$ Cannot tell $○$ No text to assess | 0.90 | 0.89 | 0.90 | 0.97 | 0.90 | 0.97 |
| 38a | [Protocol] Does the manuscript describe the process for making amendments to the trial protocol after trial commencement? | $○$ Yes $○$ No $○$ Cannot tell | 0.95 | 0.95 | 0.95 | 0.95 | 0.95 | 0.95 |
| 38b | [Protocol] Does the manuscript mention that important protocol modifications will be reported to a relevant party (e.g., REC/IRBs, trial registries, regulatory agencies)? | $○$ Yes $○$ No $○$ Cannot tell | 1.00 | 1.00 | 1.00 | 1.00 | 1.00 | 1.00 |
| 38c | [Results] Does the manuscript mention any changes made to the protocol after trial commencement? | $○$ Yes $○$ No $○$ Cannot tell $○$ No text to assess | 0.95 | 0.94 | 0.95 | 0.95 | 0.95 | 0.95 |
| 38d | [Results, if 38c=Yes] Does the manuscript mention the reasons for making one or more of those changes? | $○$ Yes $○$ No $○$ Cannot tell $○$ No text to assess $○$ Not applicable | 0.95 | 0.94 | 0.95 | 0.97 | 0.95 | 0.97 |
| 39a | Does the manuscript mention any measures to protect participant confidentiality? Check all that apply. | □ Creation of coded, de-personalized data □ Maintenance of the data and the linking code in separate locations □ Use of encrypted digital files within password-protected folders and storage media □ Limiting access to the minimum number of individuals □ Confidentiality preserved when the data are transmitted □ Other □ None reported | 0.77 | 0.89 | 0.74 | 0.94 | 0.77 | 0.94 |
| 40a | [Protocol] Does the manuscript mention who will have access to the final dataset within the research team? | $○$ Yes $○$ No $○$ Cannot tell | 0.95 | 0.93 | 0.95 | 0.95 | 0.95 | 0.95 |
| 40b | [Protocol] Does the manuscript mention whether there will be any restrictions in final dataset access within the research team? | $○$ Yes $○$ No $○$ Cannot tell $○$ Not reported | 0.90 | 0.89 | 0.90 | 0.97 | 0.90 | 0.97 |
| 41a | According to the manuscript, will trial results be disseminated, such as presenting in conferences, publishing in peer-reviewed journals, sharing with the study participants? | $○$ Yes $○$ No $○$ Not reported | 0.97 | 0.97 | 0.97 | 0.97 | 0.97 | 0.97 |
| 41b | Does the manuscript provide a timeframe for the dissemination of trial results (e.g., submitting result papers to journals within 1 year after the trial completion)? | $○$ Yes $○$ No $○$ Cannot tell | 0.97 | 0.97 | 0.97 | 0.97 | 0.97 | 0.97 |
| 41c | Does the manuscript mention if there will be/were any restrictions imposed on the result publication? | $○$ Yes $○$ No $○$ Cannot tell | 1.00 | 1.00 | 1.00 |  | 1.00 |  |
| 42a | [Protocol] Does the manuscript mention the authorship eligibility criteria for the result reports? | $○$ Yes $○$ No $○$ Cannot tell | 0.95 | 0.94 | 0.95 | 0.95 | 0.95 | 0.95 |
| 42b | Does the manuscript mention if professional writers will be/were hired? | $○$ Yes $○$ No $○$ Cannot tell | 0.97 | 0.97 | 0.97 | 0.97 | 0.97 | 0.97 |
| 43a | Does the manuscript mention the following information regarding the public access to the trial materials such as the protocol and informed consent form? Check all that apply. | □ State that they can be shared and access methods are given □ State that they can be shared but no access method is given □ State that they are not shared □ Provide a timeframe for such access, if they are not yet available □ None of the above □ Not reported | 0.93 | 0.97 | 0.75 | 0.88 | 0.96 | 0.99 |
| 44a | Does the manuscript mention the following information regarding the public access to the participant-level data? Check all that apply. | □ State that they can be shared and access methods are given □ State that they can be shared but no access method is given □ State that they are not shared □ Provide a timeframe for such access, if they are not yet available □ None of the above □ Not reported | 0.93 | 0.97 | 0.96 | 0.97 | 0.95 | 0.99 |
| 45a | Does the manuscript mention the following information regarding the public access to the analysis code? Check all that apply. | □ State that they can be shared and access methods are given □ State that they can be shared but no access method is given □ State that they are not shared □ Provide a timeframe for such access, if they are not yet available □ None of the above □ Not reported | 1.00 | 1.00 | 1.00 | 1.00 | 1.00 | 1.00 |
| 46a | According to the manuscript mention, are biological specimens collected? | $○$ Yes $○$ No $○$ Not reported | 0.88 | 0.85 | 0.88 | 0.88 | 0.88 | 0.88 |
| 46b | [If 46a=Yes] Does the manuscript describe any of the following? Check all that apply. | □ Specimen collection process □ Specimen storage, including the location of repositories □ Specimen evaluation □ Whether collected samples and associated participant-related data will be de-identified □ Other □ None reported □ No text to assess □ Not applicable | 0.80 | 0.94 | 0.96 | 0.88 | 0.60 | 0.90 |
| 47a | [Results] Given the available information from texts and diagrams/tables, if either or both are present, does the manuscript report any of the following for the participant flow? Check all that apply. | □ Number assessed for eligibility/screened □ Number eligible □ Number randomized □ Number received intended treatment □ Number analyzed for primary outcome □ Number of losses and exclusions after randomization □ Reasons for losses and exclusions after randomization □ Other □ None reported □ Supplementary figures/tables | 0.73 | 0.65 | 0.73 | 0.90 | 0.79 | 0.67 |
| 47b | [Results] Does the manuscript depict the participant flow in a diagram? | $○$ Yes $○$ No | 1.00 | 1.00 | 1.00 | 1.00 | 1.00 | 1.00 |
| 48a | Does the manuscript report the following for the recruitment process? Check all that apply. | □ Start date □ End date □ Duration □ Can’t tell □ None reported | 0.84 | 0.81 | 0.86 | 0.89 | 0.84 | 0.92 |
| 49a | Does the manuscript report the following for the follow-up process? Check all that apply. | □ Start date □ End date □ Duration □ Can’t tell □ None reported | 0.71 | 0.83 | 0.53 | 0.84 | 0.49 | 0.85 |
| 50a | [Results] Does the manuscript report that the trial was stopped early? | $○$ Yes $○$ No $○$ No text to assess | 0.90 | 0.88 | 0.90 | 0.90 | 0.90 | 0.90 |
| 50b | [Results, if 50a=Yes] Does the manuscript report the following information on early stop? Check all that apply. | □ Why the trial was stopped □ Who recommended/made the decision to stop the trial □ Other □ None reported □ No text to assess □ Not applicable | 0.91 | 0.96 | 0.83 | 0.89 | 0.96 | 0.98 |
| 51a | [Results] Given the available information from texts and tables, if either or both are present, does the manuscript report baseline characteristics by each study group? | $○$ Yes $○$ No | 0.80 | 0.76 | 0.80 | 0.80 | 0.80 | 0.80 |
| 52a | [Results] Does the manuscript report the number of participants included in the intention-to-treat, per-protocol, or as-treated analysis (overall, in each group, or both overall and in each group)? | $○$ Yes $○$ No $○$ Cannot tell | 0.75 | 0.68 | 0.75 | 0.75 | 0.75 | 0.75 |
| 53a | [Results] For any of the outcomes, does the manuscript report a summary of the outcome in each group? | $○$ Yes $○$ No | 1.00 | 1.00 | 1.00 |  | 1.00 |  |
| 53b | [Results] For any of the outcomes, does the manuscript report the estimated effect size between groups? | $○$ Yes $○$ No | 0.85 | 0.73 | 0.85 | 0.85 | 0.85 | 0.85 |
| 53c | [Results] For any of the outcomes, does the manuscript report the precision around the estimated effect size? | $○$ Yes $○$ No | 0.60 | 0.36 | 0.60 | 0.60 | 0.60 | 0.60 |
| 54a | [Results] Does the manuscript report both the absolute and relative effect size for any of the binary outcomes? | $○$ Yes $○$ No $○$ Not applicable $○$ No text to assess | 0.60 | 0.44 | 0.60 | 0.80 | 0.60 | 0.80 |
| 55a | [Results] Does the manuscript report any results from subgroup analyses? | $○$ Yes $○$ No $○$ Cannot tell | 0.75 | 0.50 | 0.75 | 0.75 | 0.75 | 0.75 |
| 55b | [Results, if 55a=Yes] When reporting the results, does the manuscript mention that the subgroup analysis is pre-specified or exploratory/post hoc? | $○$ Yes $○$ No $○$ Cannot tell $○$ No text to assess $○$ Not applicable | 0.15 | -0.05 | 0.15 | 0.79 | 0.15 | 0.79 |
| 55c | [Results] Does the manuscript report any results from adjusted analyses? | $○$ Yes $○$ No $○$ Cannot tell | 0.60 | 0.46 | 0.60 | 0.80 | 0.60 | 0.80 |
| 55d | [Results, if 55c=Yes] When reporting the results, does the manuscript mention that the adjusted analysis is pre-specified or exploratory/post hoc? | $○$ Yes $○$ No $○$ Cannot tell $○$ No text to assess $○$ Not applicable | 0.35 | 0.21 | 0.35 | 0.84 | 0.35 | 0.84 |
| 56a | [Results] Does the manuscript report any results for non-systematically assessed harms? | $○$ Yes $○$ No $○$ Cannot tell | 0.80 | 0.77 | 0.80 | 0.90 | 0.80 | 0.90 |
| 56b | [Results] Does the manuscript report the following results for harms? Check all that apply. | □ Absolute effect size □ Relative effect size □ Proportion/incidence of people experiencing an event □ Number of events per unit of time at risk □ Occurrence or non-occurrence of serious harms □ Zero counts of prespecified and systematically assessed harms □ Other □ None reported □ Supplementary figures/tables | 0.78 | 0.91 | 0.89 | 0.94 | 0.83 | 0.98 |

For conditional questions, to isolate errors unrelated to mistakes on the parent questions, we also report performance when the parent questions were answered (Table S2).

**Table S2**. Conditional questions and LLM performance. PPV: positive predictive value; NPV: negative predictive value.

| **ID** | **Question** | **Options** | **F_1_** | **GWET** | **Sensitivity** | **Specificity** | **PPV** | **NPV** |
| --- | --- | --- | --- | --- | --- | --- | --- | --- |
| 2b | [If 2a=Yes] Does the manuscript mention whether the funder is involved in the following? \|; Study design \|; Data collection, management, analysis, or interpretation \|; Writing of the report \|; Decision to submit the report for publication \| | Not reported \| Reported “involved” \| Reported “not involved”; $○$ \| $○$ \| $○$; $○$ \| $○$ \| $○$; $○$ \| $○$ \| $○$; $○$ \| $○$ \| $○$ | 0.93 | 0.90 | 0.93 | 0.94 | 0.93 | 0.94 |
| 2c | [If 2a=Yes] Does the manuscript mention any non-financial (e.g., equipment, drugs, services) support? | $○$ Yes $○$ No $○$ Not reported $○$ No text to assess | 0.85 | 0.83 | 0.85 | 0.95 | 0.85 | 0.95 |
| 3b | [If 3a=Yes] Does the manuscript mention the role of the sponsor in the study (e.g., involvement in the study design, data collection, management, analysis, and interpretation, writing of the report, decision to submit the report for publication, etc.)? | $○$ Yes $○$ No | 1.00 | 1.00 | 1.00 | nan | 1.00 | nan |
| 14d | [Protocol] If "No" to at least one of 14a,14b, and 14c, does the manuscript mention a rationale to justify why there is no provision for ancillary care, post-trial care, or compensation for trial-related harms? | $○$ Yes $○$ No $○$ Cannot tell $○$ No text to assess $○$ Not applicable | 0.75 | 0.68 | 0.75 | 0.75 | 0.75 | 0.75 |
| 15b | [If 15a=Yes] For the primary outcome or the first-mentioned primary outcome if there is more than one primary outcome, does the manuscript report the following for that outcome? Check all that apply. | □ Specific measurement □ Analysis metric □ Method of aggregation □ Measurement time point of interest □ None reported □ No text to assess | 0.83 | 0.81 | 0.84 | 0.87 | 0.77 | 0.93 |
| 15c | [If 15a=Yes] For the primary outcome or the first-mentioned primary outcome if there is more than one primary outcome, does the manuscript mention the rationale of choosing that outcome? | $○$ Yes $○$ No $○$ No text to assess | 0.75 | 0.64 | 0.75 | 0.88 | 0.75 | 0.88 |
| 15e | [If 15d=Yes] For the first-mentioned non-primary outcomes, does the manuscript report the following for that outcome? Check all that apply. | □ Specific measurement □ Analysis metric □ Method of aggregation □ Measurement time point of interest □ None reported □ No text to assess | 0.62 | 0.74 | 0.69 | 0.82 | 0.54 | 0.96 |
| 15f | [If 15d=Yes] For the first-mentioned non-primary outcomes, does the manuscript mention the rationale of choosing that outcome? | $○$ Yes $○$ No $○$ No text to assess | 0.78 | 0.69 | 0.78 | 0.89 | 0.78 | 0.89 |
| 16b | [Results, if 16a=Yes] What changes are reported? Check all that apply. | □ Change in the way that an outcome is assessed □ Original outcome replaced with new outcome □ Original outcome dropped □ New outcome added □ Change in the designation of outcomes as primary or secondary □ Other □ Can’t tell □ No text to assess □ Not applicable | 0.90 | 0.97 | 1.00 | 1.00 | 0.90 | 0.99 |
| 16c | [Results, if 16a=Yes] Does the manuscript mention the rationale for one or more of the changes? | $○$ Yes $○$ No $○$ Cannot tell $○$ No text to assess $○$ Not applicable | 0.90 | 0.89 | 0.90 | 0.90 | 0.90 | 0.90 |
| 18b | [If 18a=Yes] Is any of the following specified? Check all that apply. | □ The outcome on which the calculation was based □ Effect size □ Justification or reference of the effect size □ Type I error or confidence interval level □ One-sided or two-sided □ Type II error or power □ Statistical test □ Any allowance made for attrition or non-compliance during the study □ Other □ None reported | 0.89 | 0.82 | 0.95 | 0.83 | 0.77 | 0.97 |
| 23b | [Protocol, if 23a=Yes] Does the manuscript mention further details about the additional consent processes? | $○$ Yes $○$ No $○$ No text to assess $○$ Not applicable | 1.00 | 1.00 | 1.00 | nan | 1.00 | nan |
| 31b | [Protocol, if 31a=Yes] Is subgroup analysis specified with further details? | $○$ Yes $○$ No $○$ Cannot tell $○$ No text to assess | 0.95 | 0.93 | 0.95 | 0.95 | 0.95 | 0.95 |
| 31c | [Results, if 31a=Yes] Does the manuscript mention whether the subgroup analysis is pre-specified or exploratory/post hoc? | $○$ Yes $○$ No $○$ Cannot tell $○$ No text to assess | 0.90 | 0.89 | 0.90 | 0.97 | 0.90 | 0.97 |
| 31e | [Protocol, if 31d=Yes] Is adjusted analysis specified with further details? | $○$ Yes $○$ No $○$ Cannot tell $○$ No text to assess | 0.75 | 0.70 | 0.75 | 0.88 | 0.75 | 0.88 |
| 31f | [Results, if 31d=Yes] Does the manuscript mention whether the adjusted analysis is pre-specified or exploratory/post hoc? | $○$ Yes $○$ No $○$ Cannot tell $○$ No text to assess | 0.65 | 0.57 | 0.65 | 0.88 | 0.65 | 0.88 |
| 32b | [If 32a=No] Does the manuscript mention the rationale for not having a data monitoring committee? | $○$ Yes $○$ No $○$ Not applicable | 0.97 | 0.96 | 0.97 | 0.97 | 0.97 | 0.97 |
| 32c | [If 32a=Yes] Does the manuscript mention the following aspects of the data monitoring committee? Check all that apply. | □ Composition or intended size and characteristics of the membership □ Roles and responsibilities □ Planned method of functioning □ Degree of independence from the sponsor and investigators □ Other □ None reported □ No text to assess □ Not applicable | 0.88 | 0.95 | 0.72 | 0.99 | 0.81 | 0.96 |
| 33b | [If 33a=Yes] Does the manuscript mention the following aspects of the interim analyses? Check all that apply. | □ Rationale for the analysis □ Statistical methods used for the analysis □ Timing of the analysis □ Who will conduct the analysis □ If the analysis will be masked □ Who will have the access to the analysis results □ Whether those with access to the results will remain masked □ Whether masking will be maintained when any adaptations to the trial are made □ Other □ None reported □ No text to assess □ Not applicable | 0.88 | 0.97 | 0.76 | 0.97 | 0.99 | 0.98 |
| 34b | [If 34a=Yes] Does the manuscript mention who has the ultimate authority to stop the trial? | $○$ Yes $○$ No $○$ No text to assess $○$ Not applicable | 0.88 | 0.86 | 0.88 | 0.96 | 0.88 | 0.96 |
| 36b | [Protocol, if 36a=Yes] Does the manuscript mention the following aspects of trial auditing? Check all that apply. | □ Procedures of auditing □ Frequency of auditing □ Personnel involved and their degree of independence from the trial investigators and sponsor □ Other □ None reported □ No text to assess □ Not applicable | 0.93 | 0.98 | 1.00 | 0.93 | 0.98 | 0.99 |
| 38d | [Results, if 38c=Yes] Does the manuscript mention the reasons for making one or more of those changes? | $○$ Yes $○$ No $○$ Cannot tell $○$ No text to assess $○$ Not applicable | 0.95 | 0.94 | 0.95 | 0.97 | 0.95 | 0.97 |
| 46b | [If 46a=Yes] Does the manuscript describe any of the following? Check all that apply. | □ Specimen collection process □ Specimen storage, including the location of repositories □ Specimen evaluation □ Whether collected samples and associated participant-related data will be de-identified □ Other □ None reported □ No text to assess □ Not applicable | 0.85 | 0.96 | 0.96 | 0.90 | 0.97 | 0.90 |
| 50b | [Results, if 50a=Yes] Does the manuscript report the following information on early stop? Check all that apply. | □ Why the trial was stopped □ Who recommended/made the decision to stop the trial □ Other □ None reported □ No text to assess □ Not applicable | 0.91 | 0.96 | 0.83 | 0.89 | 0.96 | 0.98 |
| 55b | [Results, if 55a=Yes] When reporting the results, does the manuscript mention that the subgroup analysis is pre-specified or exploratory/post hoc? | $○$ Yes $○$ No $○$ Cannot tell $○$ No text to assess $○$ Not applicable | 0.35 | 0.20 | 0.35 | 0.84 | 0.35 | 0.84 |
| 55d | [Results, if 55c=Yes] When reporting the results, does the manuscript mention that the adjusted analysis is pre-specified or exploratory/post hoc? | $○$ Yes $○$ No $○$ Cannot tell $○$ No text to assess $○$ Not applicable | 0.50 | 0.39 | 0.50 | 0.88 | 0.50 | 0.88 |

### **Section C. Additional Model Performance Analyses**

*Results by question type and relevant guideline*

The performance breakdown by question type (check-one vs. check-all-that-apply) and relevant guideline (SPIRIT vs. CONSORT vs. both) for the best-performing model is provided in Table S3. The model performs better on check-one questions compared to check-all-that-apply questions in terms of F_1_ (0.829 vs. 0.805) and sensitivity (0.829 vs. 0.814). While this is as expected, because check-one questions tend to be less complex than check-all-that-apply, the difference was small, suggesting that our automated approach handles increased question complexity reasonably well. At the same time, performance in terms of specificity and Gwet’s AC1 is higher for check-all-that-apply questions because these metrics account for agreement on unselected options that are more common in multiple-choice settings.

|  | F_1_ (95% CI) | Sensitivity (95% CI) | Specificity (95% CI) | Gwet’s AC1 (95% CI) |
| --- | --- | --- | --- | --- |
| *Question type* |  |  |  |  |
| Check-one | 0.829 (0.794-0.861) | 0.829 (0.794–0.861) | 0.883 (0.863–0.901) | 0.771 (0.724-0.815) |
| Check-all-that-apply | 0.805 (0.759-0.847) | 0.814 (0.766–0.859) | 0.903 (0.878–0.925) | 0.855 (0.815-0.893) |
| *Applicable guideline* |  |  |  |  |
| SPIRIT | 0.840 (0.813-0.865) | 0.842 (0.816–0.868) | 0.899 (0.883–0.915) | 0.819 (0.786-0.852) |
| CONSORT | 0.780 (0.742-0.815) | 0.786 (0.748–0.821) | 0.871 (0.852–0.890) | 0.736 (0.688-0.782) |

**Table S3**. GPT-5 performance on 119 questions with spirit-consort-tm-identified snippets and hand-crafted prompt templates, grouped by question type and corresponding reporting guidelines. CI: confidence interval.

Regarding reporting guidelines, the model performs better on SPIRIT-related questions compared with CONSORT-related questions (F_1_: 0.840 vs. 0.780). The performance gap was larger between SPIRIT-specific and CONSORT-specific questions (F_1_: 0.884 vs. 0.750). One possible explanation is that CONSORT-specific questions often require assessing nuanced interpretations of trial results (10 of the 24 CONSORT-specific questions concern results) which are typically diffuse and lengthy in publications and might be presented in figures and tables that the current approach does not address. This may require deeper contextual reasoning and make it harder for the LLM to identify the most relevant information within larger text spans. In contrast, SPIRIT-specific questions tend to be more focused, with shorter supporting passages, and are therefore likely easier to answer. This difference is also reflected in the length of text snippets associated with each question: the average snippet length was approximately 68 tokens for SPIRIT-specific questions, compared with approximately 195 tokens for CONSORT-specific questions.

LLMs tended to underperform on questions requiring more detailed information about specific elements (e.g., “For the first-mentioned non-primary outcomes, does the manuscript mention the rationale of choosing that outcome?”), while performing well on questions that ask whether the manuscript explicitly mentions a given element (e.g., “Does the manuscript mention the plans for post-trial care?”).

*Results by prompting strategy*

To assess the contribution of prompting strategies, we compared the default approach (hand-crafted prompt template with chain-of-though [CoT] reasoning) to two alternatives: the same template without CoT and an LLM-generated prompt template. The results are shown in Table S4. The hand-crafted template performs slightly better than the LLM-generated template in guiding GPT-5 to perform the task, although the improvements are modest (+0.8 pp F_1_, +0.8 pp in sensitivity, +0.4 pp in specificity, and +0.8 pp Gwet’s AC1). Removing CoT leads to a slightly larger performance drop (-1.0 pp F_1_, -1.2 pp sensitivity, -0.3 pp specificity, and -1.4 pp Gwet’s AC1).

| Prompt template | F_1_ (95% CI) | Sensitivity (95% CI) | Specificity (95% CI) | Gwet’s AC1 (95% CI) |
| --- | --- | --- | --- | --- |
| Hand-crafted prompt with CoT (Default) | 0.822 (0.794–0.847) | 0.824 (0.796–0.851) | 0.889 (0.874–0.904) | 0.796 (0.760–0.829) |
| Hand-crafted prompt w/o CoT | 0.812 (0.783–0.839) | 0.812 (0.783–0.839) | 0.886 (0.869–0.901) | 0.782 (0.743–0.818) |
| LLM-generated prompt | 0.814 (0.787–0.84) | 0.816 (0.788–0.842) | 0.885 (0.869–0.901) | 0.788 (0.751–0.822) |

**Table S4**. LLM performance on 119 questions given spirit-consort-tm-identified relevant text and different constructions of question answer instruction template. CoT prompting is included unless specified. CI: confidence interval.

Given the additional cost and longer response time associated with CoT-style reasoning (Section D), its practical utility therefore appears limited. One likely explanation is that the presence of specific words and phrases in retrieved text snippets is often sufficient to answer the questions, reducing the need for complex reasoning.

We focused on generating question-specific instructions using a well-designed prompt template. The results suggest that GPT-5, in particular, can reliably generate accurate instructions using this shared prompting framework, although model performance on individual questions could be further improved through targeted question-specific prompt optimization. Given the substantial time and labor involved in manual assessment, developing detailed and question-specific instructions may be a feasible approach for further improving model performance.

We examined the LLM-generated prompts for each question and the reasoning traces produced in model responses. Overall, the LLM-generated prompts for each question were substantially longer than our hand-crafted prompts and frequently included “guidance on options”, describing what information should be identified in the provided text snippet. However, these explanations were often superficial and did not meaningfully support reasoning in the RCT reporting context. For example, for the question “Does the manuscript specify one or more outcomes as the non-primary outcome?”, the model generated the guidance: “Choose this if the manuscript explicitly identifies one or more outcomes that are not primary (e.g., secondary, other, exploratory)”. This instruction largely repeats the information provided rather than clarifying how to interpret or detect such information in practice.

In contrast, the reasoning traces generated by the model illustrate how specific textual evidence is connected to the final selections. For instance, the sentence “Placebos were made with the same taste and appearance but without the principal ingredients that present in the red ginseng extract. . . ” led GPT-5 to infer that “the interventions are made the same in appearance and flavor/taste, with no mention of number, timing, or duration,” and therefore to select *appearance* and *flavor/taste* as the appropriate options. Overall, these observations suggest that CoT prompting improves transparency by making explicit how textual evidence leads to specific reporting assessments, while it has limited impact on performance.

### **Section D. Cost and computational efficiency**

We ran Qwen-2.5 locally, avoiding per-article API costs, while GPT-5 incurred an estimated cost of approximately $1.45 per article. However, local deployment requires additional computational resources, with Qwen-2.5 requiring longer inference time than GPT-5 (5.33 vs. 2.35 minutes per article) and incurring infrastructure and electricity costs.

Evidence retrieval incurs additional computation costs compared with using expert-annotated snippets, increasing cost ($1.45 vs. $1.09 per articles) and inference time (2.35 vs. 1.89 minutes per articles), indicating that expert-annotated snippets are typically shorter than the snippets retrieved by the spirit-consort-tm model, resulting in shorter processing time.

Incorporating CoT reasoning into the prompt increased the response time by more than fivefold (0.44 vs. 2.35 minutes per article) and cost by more than threefold ($0.41 vs. $1.45 per article). On the other hand, using the LLM-generated prompts instead of the hand-crafted prompts leads to shorter processing time while the cost is similar to that of the default hand-crafted prompts.

In-context examples incur additional inference time and cost, especially when the selection of similar examples relies on embedding-based semantic similarity ($1.94 and 2.39 minutes with spirit-consort-tm embeddings; $1.86 and 6.49 minutes with Qwen-8B embeddings) . Finding similar examples using Qwen-8B embeddings is substantially slower because the model is run locally.
